# Prognostic value of an integrated human papilloma virus and immunoscore model to predict survival in vulva squamous cell carcinoma

**DOI:** 10.1101/2024.09.11.24313475

**Authors:** Rammah Elnour, Ingjerd Helstrup Hindenes, Malene Færevaag, Ingrid Benedicte Moss Kolseth, Liv Cecilie Vestrheim Thomsen, Anne Christine Johannessen, Daniela Elena Costea, Line Bjørge, Harsh Nitin Dongre

**Affiliations:** Center for Cancer Biomarkers CCBIO and Gade Laboratory of Pathology, Department of Clinical Medicine, University of Bergen, Bergen, Norway; Department of Chemistry, University of Bergen, Bergen, Norway; Department of Obstetrics and Gynecology, Haukeland University Hospital, Bergen, Norway; Center for Cancer Biomarkers CCBIO, Department of Clinical Science, University of Bergen, Bergen, Norway; Department of Pathology, Laboratory Clinic, Haukeland University Hospital, Bergen, Norway

**Keywords:** immunoscore, prognostic marker, vulva squamous cell carcinoma, immune-oncology, digital pathology

## Abstract

**Background:** While the prognostic value of immune-related biomarkers is well characterized in many solid tumors, their significance in vulva squamous cell carcinoma (VSCC) remains unclear. Here, we report a comprehensive analysis of programmed death-ligand 1 (PD-L1) and immune cell infiltrates in VSCC and establish immunoscore models for classification of the disease.

**Methods:** Archival tissues, immunohistochemistry, and digital quantification were used to investigate the number of CD4+, CD8+, CD68+, CD14+, FoxP3+, and PD-L1+ cells in epithelial and stromal compartments of VSCC (*n*=117). Immunoscores were developed using these parameters and applying the least absolute shrinkage and selection operator (LASSO) to identify predictors of survival. Immunoscores were integrated with HPV status, as determined by mRNA *in situ* hybridization, to construct internally validated nomograms. The models were assessed using Harrell’s concordance-index (c-index), calibration plots, Kaplan-Meier curves, and decision curve analysis.

**Results:** Advanced VSCC (FIGO stage III/IV) was characterized by high numbers of CD68+ macrophages and PD-L1+ cells (Spearmans’ correlation, ρ>0.80) in the epithelium. PD-L1 status independently predicted poor progression free survival (PFS) (HR=1.80, (95% CI (1.024-3.170), p=0.041). High stromal CD68+ or CD14+ myeloid cell infiltration was associated with poor PFS and disease specific survival (DSS) (p<0.05). Immunological parameters were used to determine immunoscores. Immunoscore^PFS^ and immunoscore^DSS^ were independent prognosticators of PFS (p=0.001) and DSS (p=0.007) respectively. Integrating immunoscores with HPV status (IS-HPV index) improved the prognostic impact of the models. The c-index of IS-HPV index^PFS^ was 0.750 for prediction of PFS compared to 0.666 for HPV status and 0.667 for immunoscore^PFS^. The c-index of IS-HPV index^DSS^ was 0.752 for predicting DSS compared to 0.631 for HPV status and 0.715 for immunoscore^DSS^.

**Conclusion:** In summary, an index based on HPV status and an immunoscore built on PD-L1 expression and immune cell infiltrates could potentially serve as a prognostic tool to refine risk stratification in VSCC. Further validation is warranted to demonstrate clinical utility.

**WHAT IS ALREADY KNOWN ON THIS TOPIC:** Immunoscores have emerged as promising biomarkers in several solid tumors, however the prognostic potential of these models in vulva squamous cell carcinoma (VSCC) remains unexplored. Immunoscores are traditionally based on quantification of CD3+ and CD8+ T lymphocytes within the tumor core and invasive margin. However recent studies have highlighted the prognostic significance of additional immune cell subtypes, such as macrophages, in predicting patient survival. Advanced quantitative methods that enable the determination of immunoscores, incorporating expression of immune checkpoint proteins and infiltration of various immune cell populations within tumors, warrant further investigation.

**WHAT THIS STUDY ADDS:** This study investigates PD-L1 expression and distinct T lymphocyte and myeloid cell subsets within the spatial architecture of VSCC to identify immunological markers with prognostic value. Computational analysis identified PD-L1+ cells, CD8+ T lymphocytes, FoxP3+ T lymphocytes, and CD14+ myeloid cells as significant survival predictors in VSCC, leading to the development of immunoscores. These immunoscores demonstrated superior predictive performance compared to the individual parameters used to construct the models. Moreover, integrating immunoscores with HPV status and constructing, for the first time, an immunoscore-HPV index nomogram enhanced the prognostic accuracy and net benefit of the models.

**HOW THIS STUDY MIGHT AFFECT RESEARCH, PRACTICE OR POLICY:** The immunoscore-HPV index proposed in this study holds potential as a rapid diagnostic tool to enhance risk stratification in VSCC and facilitate the identification of patients who may benefit from less aggressive treatment approaches. Incorporating immunoscores with clinicopathological or molecular features linked to carcinogenesis in prognostic models could improve classification strategies in VSCC and other cancer types.

## Introduction

Vulvar squamous cell carcinoma (VSCC) is a relatively rare but heterogenous disease, with 47,300 new cases and 18,600 deaths globally each year (1). VSCC accounts for six percent of all gynecological malignancies (2), and its incidence rates have been increasing steadily (3). Despite recent surgical advances, 12-37% of VSCC patients experience disease recurrence, with a five-year survival rate ranging between 30-60% (4, 5). For metastatic disease, the five-year survival rate is reported to be below 30% (6). Beyond surgical resection and radiochemotherapy, treatment options are limited, especially for managing advanced stage disease (7). While precision medicine guided by molecular subtyping has been adopted in various cancers, efforts to incorporate molecular risk stratification in VSCC have only recently begun. VSCC is characterized by two distinct etiopathogenic pathways: high risk human papilloma virus (HPV)-associated VSCC, driven by the integration of the HPV genome and the expression of the E6/E7 oncoproteins, and HPV-independent VSCC which often arises in the context of inflammatory vulvar lesions such as lichen sclerosus (8, 9). Studies have shown that HPV-associated VSCC has better prognosis compared to the more common HPV-independent VSCC (10, 11). Thus, identifying biomarkers to predict survival especially in HPV-independent VSCC is urgently needed.

The cancer immune contexture, encompassing the infiltration of various immune cell populations and the expression of immune checkpoint proteins (12), is currently being analyzed to identify biomarkers with predictive and prognostic potential in several gynecological cancers (13). Targeting programmed cell death protein-1 (PD-1) with an immune checkpoint inhibitor, has been associated with prolonged survival in a subset of patients with advanced, recurrent, or metastatic VSCC (14). This suggests that modulating the tumor immune microenvironment (TIME) is a viable therapeutical strategy with significant impact on survival, however immunotherapy has not yet been included into standard treatment regimens. Furthermore, the prognostic value of immune checkpoints and specific immune cell subsets within the spatial architecture of VSCC remains unclear (15-18). Recently, studies have indicated that the TIME can be leveraged to establish cancer classification strategies with clinical utility. Indeed, the immunoscore became the first standardized consensus assay for classifying hot and cold tumors based on the infiltration of CD3+ and CD8+ T lymphocytes in the tumor core and invasive margin in colon cancer (19-21). There have only been a few studies focused on establishing prognostic models, such as nomograms, capable of stratifying VSCC patients and predicting survival. To our knowledge, immunoscore models based on spatially resolved immunological features have yet to be reported for this disease. Developing prognostic tools based on compartment-specific infiltration patterns of immune cell subsets in both HPV-associated and HPV-independent tumors may enhance personalized medicine and allow for individual risk stratification of VSCC.

In this study, we report a comprehensive *in-situ* analysis of PD-L1 expression and distinct T lymphocyte- and myeloid cell populations in VSCC, revealing components of the immune landscape associated with disease progression. Using these immunological features, we established a novel immunoscore model for VSCC. Further, we integrated the immunoscore with HPV status and assessed the prognostic value of the resulting immunoscore-HPV status index in predicting risk progression in VSCC.

## Materials and Methods

### Study population

One hundred and thirty-eight patients diagnosed with VSCC and undergoing treatment at Haukeland University Hospital (HUS), Bergen, Norway between 1999-2017 were included in this retrospective study. The patients were treatment naïve, prospectively sampled, and staged according to the FIGO (International Federation of Gynecology and Obstetrics) criteria. Written consent was obtained from all participants prior to inclusion. Most of the patients were registered in Bergen Gynecologic Cancer Biobank (GYNCAN). A few tumor samples were obtained from the diagnostic biobank at the Department of Pathology (HUS). Cases with missing sample, insufficient epithelial and/or stromal tissue material, neoadjuvant treatment, or withdrawn consent were excluded from this study (*n* = 21). Clinicopathological data were extracted from medical journals as previously described (10). This study was approved by the Regional Committee for Medical and Health Research Ethics Norway (REK Nos: 2017/279 and 2014/1907).

### Sample collection and tissue microarray construction

Archival formalin-fixed paraffin embedded (FFPE) sections were hematoxylin and eosin stained to select the areas for construction of tissue microarrays (TMA). A minimum of two 1 mm cylinder biopsies from the tumor front areas of primary tumors, recurrences, and metastases were punched out and placed in paraffin blocks using a precision instrument (Beecher Instruments, Silver Spring, MD, USA). TMA sections of 4-μm thickness were cut using a microtome (Leica Microsystems, Wetzlar, Germany) and mounted on Superfrost Plus glass slides (Thermo Fisher Scientific, Waltham, MA, USA). Slides were then incubated at 56°C for 24 hours and stored at 4°C in preparation for immunostaining.

### Immunohistochemistry and HPV detection

Immunohistochemistry (IHC) was performed on TMA sections using a Ventana Benchmark ULTRA Autostainer (Roche Tissue Diagnostics, Tucson, AZ, USA), according to the manufacturer’s instruction. CD4+ helper T cells, CD8+ cytotoxic T cells, CD68+ macrophages, CD14+ monocytes, FoxP3+ regulatory T cells, and PD-L1+ cells were identified using the antibodies summarized in Supplementary Table 1. The OptiView DAB IHC detection kit (Roche Tissue Diagnostics) was used for detection of primary antibodies and visualization. The detailed protocols for p16 and p53 IHC, HPV DNA PCR, and HPV mRNA *in situ* hybridization (ISH) are described elsewhere (10, 22).

### Digital quantification and evaluation

Immunostained TMAs were digitally scanned using a Hamamatsu NanoZoomer-XR whole slide scanner (Hamamatsu Photonics, Hamamatsu City, Japan) at 40x magnification and analyzed using the open-source software QuPath (version 0.4.0) (23). For digital quantification, epithelial and stromal compartments of TMA cores were manually annotated using QuPath tools (Supplementary Figure 1A). QuPath automated algorithms for setting vector stains and color-deconvolution were then used to detect cell nuclei and immune markers based on adjusted hematoxylin and 3, 3’-diaminobenzidine (DAB) thresholds (Supplementary Figure 1B-C). The hematoxylin and DAB thresholds were maintained between slides of each marker to acquire homogenous results. Routine calibration was performed together with a qualified pathologist (DEC). The number of positive cells for each marker per mm^2^ compartment was then calculated and the mean of all cores from each individual patient sample was used for analysis. CD4+, CD8+, CD68+, CD14+, and FoxP3+ immune cells were dichotomized into high and low infiltration based on median positive cell counts per mm^2^ epithelial or stromal compartment for each respective marker (Supplementary Figure 2). PD-L1 status was determined by using both the tumor proportion score method (TPS: number of PD-L1+ tumor cells divided by the total number of tumor cells multiplied by 100) and the combined positive score method (CPS: total number of PD-L1+ tumor cells and PD-L1+ stromal cells divided by the total number of tumor cells multiplied by 100) (24). Cases with a discriminative cut-off value of TPS > 1 or CPS > 1 were considered PD-L1 positive.

### Statistical analysis

All statistical analysis and data visualization was performed using IBM SPSS 25.0 (SPSS, Chicago, IL) and R (version 4.3.0 or higher). Differences concerning infiltration of cells within epithelial or stromal compartments were investigated using the Wilcoxon-Signed Rank Test. The chi-squared test was used to explore associations between immune subsets and clinicopathological features. The Mann-Whitney U test was used to study differences in the number of infiltrating cells based on clinicopathological features. Correlations between different cell populations were analyzed using the Spearmans’ rank correlation coefficient. Survival analysis was performed using the R packages “survival” (v.3.5-7) and “survminer” (v.0.4.9) for Kaplan-Meier estimator, log-rank test, and Cox regression models. p-values ≤ 0.05 were considered statistically significant. Parameters that were significant in univariate analysis were further analyzed in multivariate analysis. Progression free survival (PFS) was calculated as the time in months from the date of termination of primary treatment to the date of recurrence or death. Disease specific survival (DSS) was calculated as the time in months from date of primary treatment to the date of death from disease. Patients alive at last contact or patients deceased from other causes were censored. The “glmnet” package (v.4.1-8) was used for the least absolute shrinkage and selection operator (LASSO) to determine immunoscores based on CD4+, CD8+, CD68+, CD14+, FoxP3+, and PD-L1+ expression in epithelial and stromal compartments of tumors. Immunoscores were dichotomized using the maximally selected rank statistics method in the survminer package or by median values. Immunoscores and HPV mRNA ISH were used to construct logistic regression models and nomograms using the “rms” (v.6.7-1) package and “dcurves” (v.0.4.0) was used for decision curve analysis.

## Results

### Advanced stage VSCC and HPV-independent tumors displayed higher expression of PD-L1 and infiltration of CD68+ macrophages

The number of infiltrating CD4+, CD8+, CD68+, CD14+, or FoxP3+ cells (Figure 1A) were higher in the stroma of VSCC compared to the epithelium (p < 0.05), however no significant difference was observed in the amount of PD-L1+ cells (Supplementary Figure 3A). CD68+ and CD14+ myeloid cells were the most prevalent immune cell subsets (Supplementary Table 2), and higher infiltration of these cells in the epithelium was linked to clinicopathological features including tumor size (Supplementary Table 3). Advanced tumors (FIGO stage III and IV) were more highly populated with epithelial PD-L1+ and CD68+ cells (Figure 1B). Accordingly, tumors larger in size (> 4cm) or with lymph node metastasis were found to have increased numbers of these subsets in the epithelium. Larger tumors also demonstrated higher infiltration of CD14+ monocytes, in both the epithelium and stroma (Supplementary Figure 3B-C). Within the advanced VSCC subgroup, HPV-independent tumors (as determined by HPV mRNA ISH) demonstrated a higher number of PD-L1+ cells in the stroma compared to HPV-associated counterparts (p < 0.05) (Figure 1C). Moreover, HPV-independent tumors demonstrated higher stromal infiltration of both CD68+ and CD14+ myeloid subsets in early stage VSCC (FIGO stage I and II). Further, the number of epithelial CD68+ macrophages and stromal CD14+ monocytes were higher in tumors with *TP53* mutations and no p16 expression (p53+/p16-) compared to the p53-/p16+ subtype (Supplementary Figure 3D-E). Among T lymphocyte subsets, CD8+ cytotoxic T cells were the predominant group, whereas the infiltration of CD4+ helper T cells was low, particularly in the epithelial compartments (Supplementary Table 2). Apart from FoxP3+ regulatory T cells, which were associated with disease specific survival (p < 0.05), no significant associations were found between infiltration of T cell subsets and clinicopathological features (Supplementary Table 3). To explore the correlations between PD-L1 and immune cell infiltration in the epithelial and stromal compartments, the Spearmans’ rank correlation coefficient was utilized. The number of PD-L1+ cells was positively correlated with all immune subsets in both compartments (p < 0.05) (Figure 1D). The strongest correlation was identified between PD-L1+ cells and CD68+ macrophages in the epithelium (ρ = 0.80) and stroma (ρ = 0.71) (Figure 1E-F) whereas CD4+, CD8+, CD14+, and FoxP3+ infiltrates were moderately correlated with PD-L1+ (ρ = 0.43-0.58).

**Figure 1:**
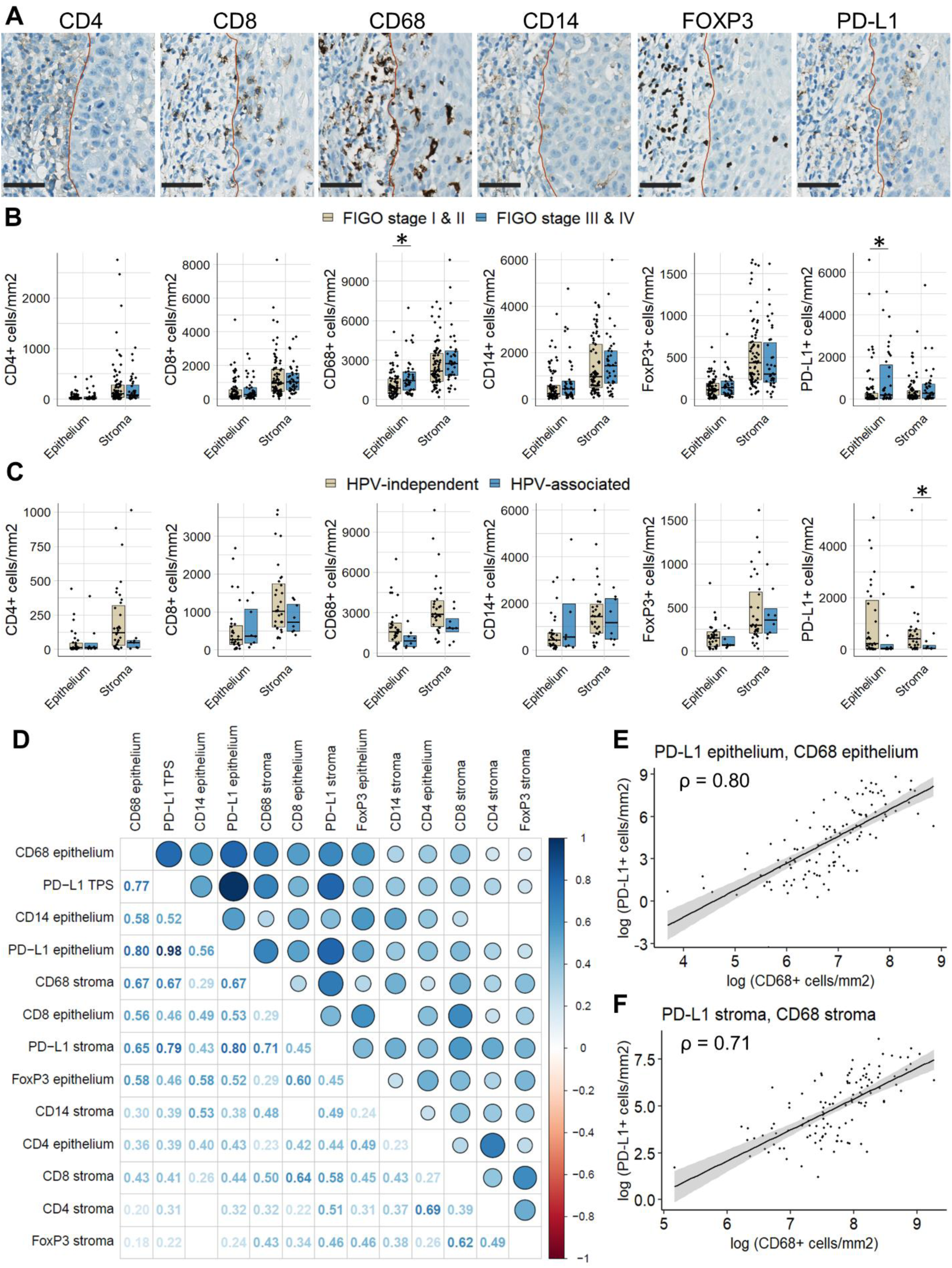
PD-L1 expression and immune cell infiltration in VSCC. (A) Representative immunohistochemical staining of CD4+, CD8+, CD68+, CD14+, FoxP3+, and PD-L1+ cells. Scale bar = 50 μm. (B) Box plots depicting differences in the number of cells per mm^2^ based on FIGO stage. (C) Box plots depicting differences in the number of cells per mm^2^ based on HPV status, as determined by HPV mRNA ISH, within the advanced VSCC subgroup (FIGO stage III/IV). p-values ≤ 0.05 were considered statistically significant. The box shows the interquartile range, and medians are highlighted with a black line. Individual cases are represented by black dots. (D) Matrix showing correlations between immune subsets. Spearmans’ rank correlation coefficients (ρ) are provided in each box of the matrix. Larger circles with darker shades of blue demonstrate stronger positive correlations. Correlations that were not statistically significant (p > 0.05) are shown as blank. Scatterplots with fitted regression lines and 95% confidence interval showing correlations between PD-L1+ and CD68+ cells in (E) epithelial and (F) stromal compartments.

### Positive PD-L1 status independently predicted poor PFS

Traditional prognostic factors for VSCC, including age and HPV status, were significantly linked to survival in this cohort (Supplementary Table 4). The analysis was expanded to assess the prognostic impact of stratifying patients by immune cell infiltration and PD-L1 status. Kaplan-Meier analysis revealed positive PD-L1 status (TPS > 1) or high stromal infiltration of CD14+ monocytes to be associated with poor PFS (log rank test: p = 0.002 and p = 0.015, respectively) (Figure 2A). This was also the case for high infiltration of CD68+ macrophages in stromal or epithelial compartments (log rank test: p = 0.019 and p = 0.042 respectively) (Figure 2A and Supplementary Figure 4A-B). Furthermore, high infiltration of either CD68+ macrophages or CD14+ monocytes in the stroma predicted poor DSS (log rank test: p = 0.011 and p = 0.027 respectively). In contrast, high epithelial infiltration of FoxP3+ T cells was associated with more favorable DSS outcomes (log rank test: p = 0.036) (Figure 2B). Results from the univariate Cox analysis aligned with these findings however, after adjusting for age and HPV status in the multivariate Cox analysis, only PD-L1 status (TPS > 1) remained an independent prognostic indicator for PFS (HR = 1.80, p = 0.041). In contrast PD-L1 status as determined by CPS > 1, was not significantly associated with PFS (Table 1). None of the investigated markers demonstrated independent prognostic value for DSS after adjusting for confounders. In this cohort, 58/117 primary tumors (49.6%), 21/29 recurrences (72.4%), and 25/36 metastatic lesions (69.4%) were PD-L1 positive (TPS > 1). Moreover, variations in PD-L1 status were evident between primary tumors and their respective recurrent or metastatic counterparts (Figure 2C-D). Within the cohort of PD-L1 negative primary tumors, eight of the matched recurrences (61.5%) and four of the matched metastases (30.8%) were PD-L1 positive respectively (Figure 2E-F). Consistent with PD-L1+ infiltration patterns, positive PD-L1 status was associated with HPV-independent VSCC and the p53+/p16-subtype (Table 2).

**Figure 2:**
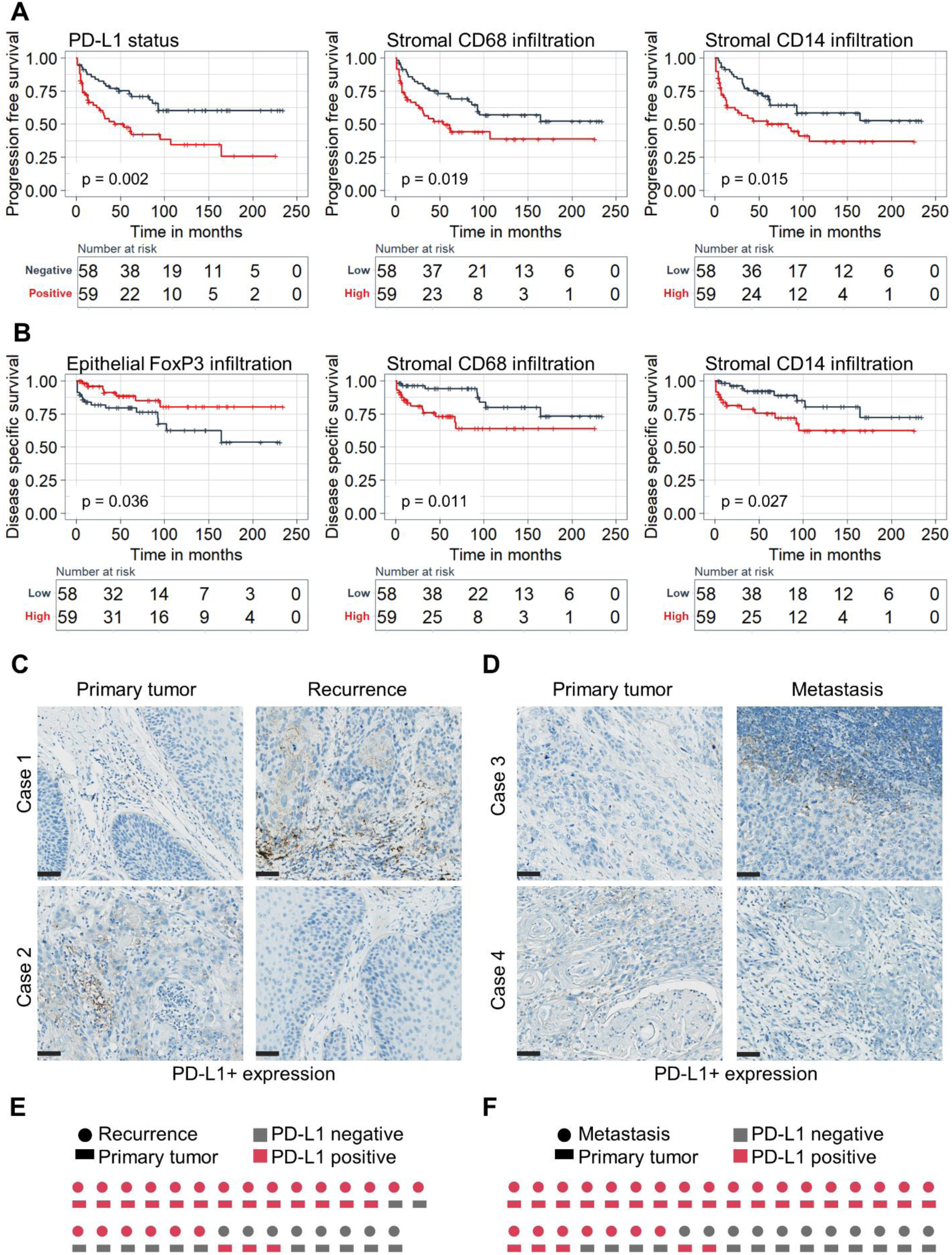
Prognostic value of PD-L1 status and immune cell infiltration. Kaplan-Meier curves demonstrating influence of PD-L1 status and stromal CD68+ or CD14+ infiltration on (A) progression free- and (B) disease specific survival. p-values < 0.05 were considered statistically significant based on the log rank test. Representative immunohistochemical staining of cases with discordant PD-L1 expression in primary tumors and corresponding (C) recurrences or (D) metastatic lesions. Scale bar = 50 μm. PD-L1 status patterns in primary tumors and matched (E) recurrences or (F) metastatic lesions.

**Table 1:**
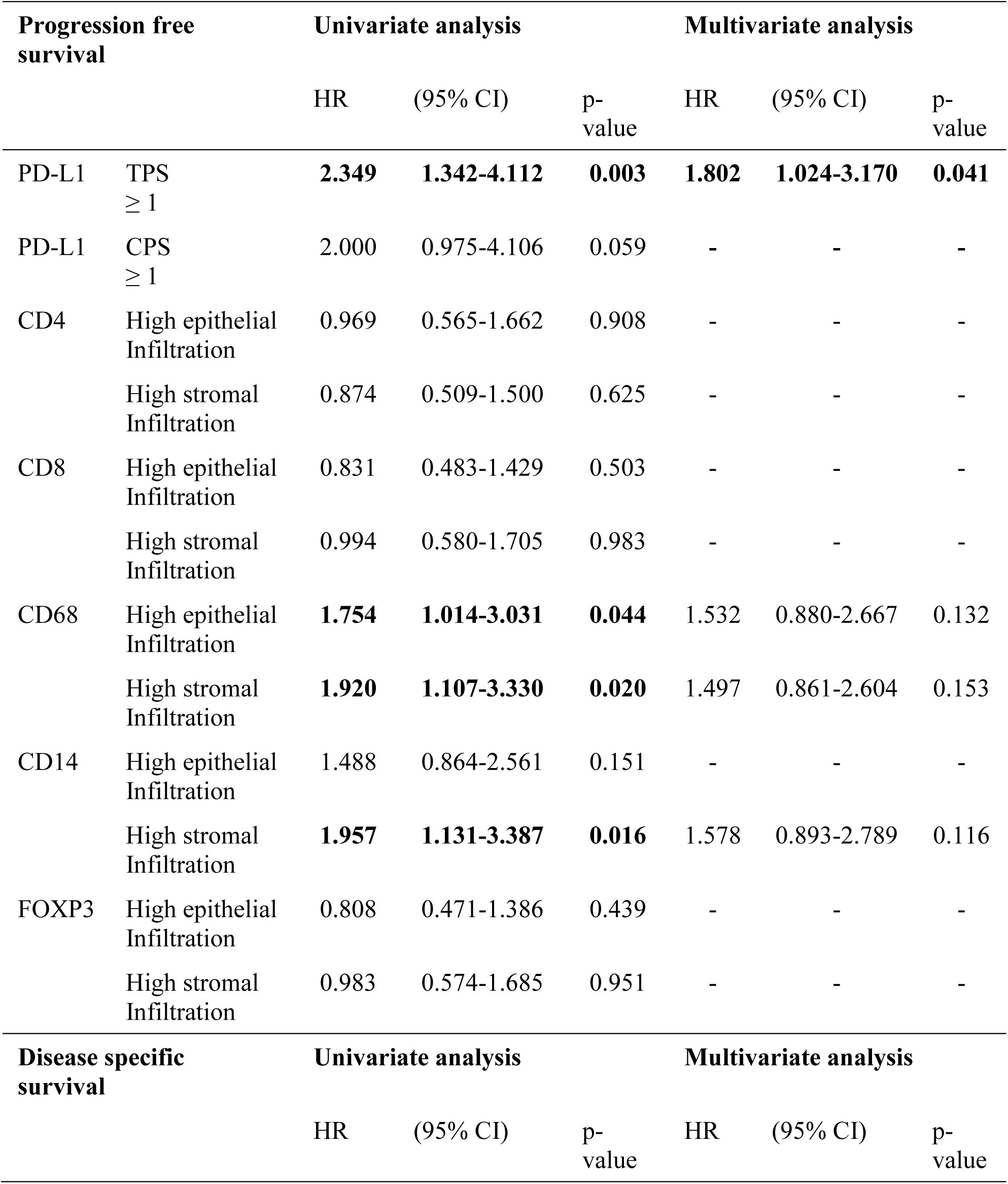
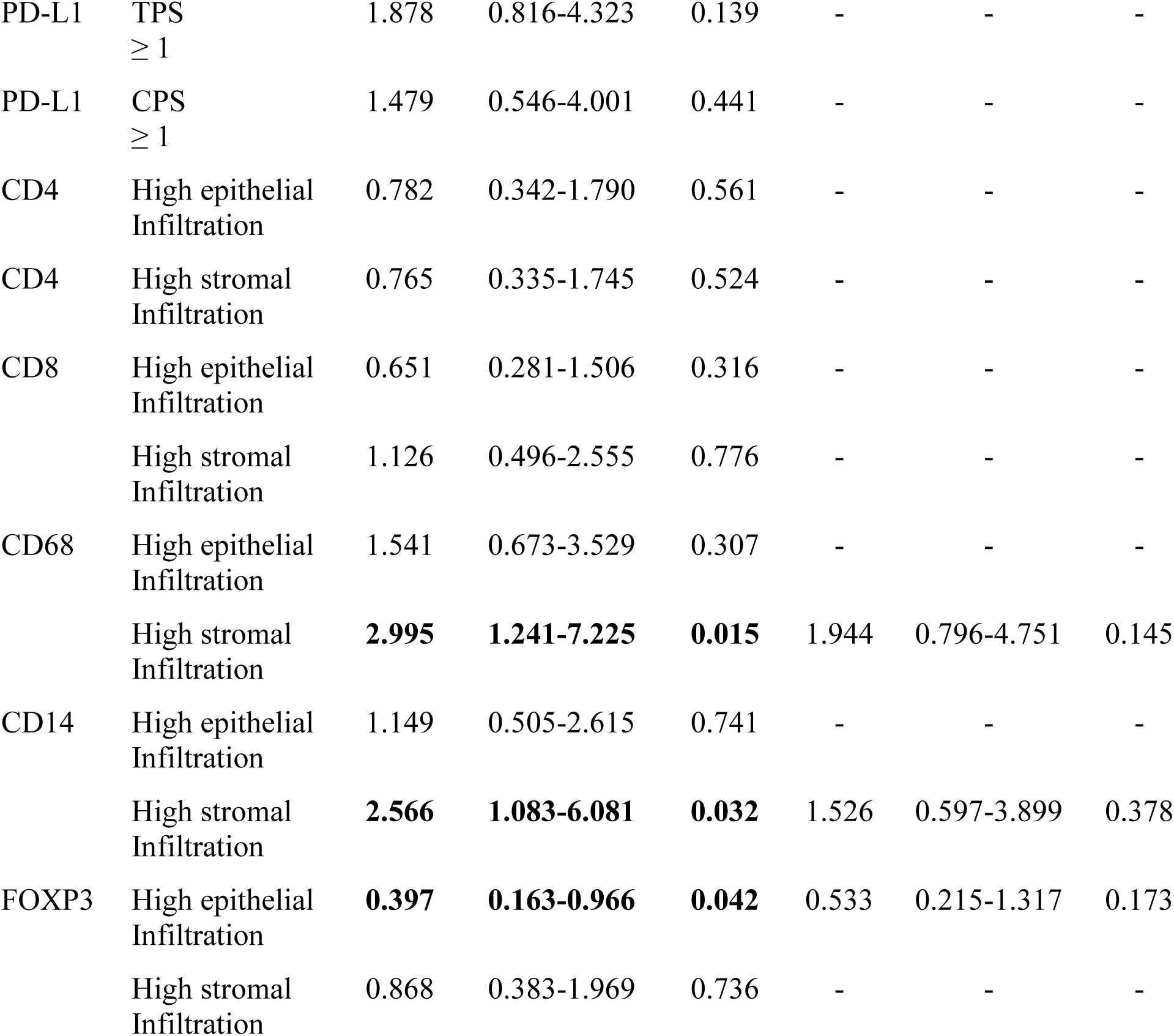
Influence of PD-L1 status and immune cell infiltration on progression free- and disease specific survival. In the multivariate analysis PFS was adjusted for age and HPV mRNA ISH. DSS was adjusted for age, tumor size, and HPV mRNA ISH.

**Table 2:** Relationship between PD-L1 status, as determined by TPS > 1, and clinicopathological features.

| Parameters | PD-L1 Status | | | $\chi^2$<br>(p-value) |
| --- | --- | --- | --- | --- |
|  | n=117 | Positive<br>(%) | Negative<br>(%) |  |
| **Age** |  |  |  |  |
| < 71 years | 57 (48.7%) | 24 (20.5%) | 33 (28.2%) | 3.079 |
| ≥ 71 years | 60 (51.3%) | 35 (29.9%) | 25 (21.4%) | (0.079) |
| **BMI** |  |  |  |  |
| Normal | 31 (26.5%) | 13 (11.1%) | 18 (15.4%) | 1.462 |
| Overweight/obese | 54 (46.2%) | 30 (25.6%) | 24 (20.5%) | (0.227) |
| Unknown | 32 (27.3%) | 15 (12.8%) | 17 (14.5%) |  |
| **FIGO stage** |  |  |  |  |
| Stage I & II | 78 (66.7%) | 35 (29.9%) | 43 (36.8%) | 2.889 |
| Stage III & IV | 39 (33.3%) | 24 (20.5%) | 15 (12.8%) | (0.089) |
| **Tumor size** |  |  |  |  |
| ≤ 4cm | 81 (69.2%) | 37 (31.6%) | 44 (37.6%) | 2.374 |
| > 4cm | 36 (30.8%) | 22 (18.8%) | 14 (12.0%) | (0.123) |
| **Lymph node metastasis** |  |  |  |  |
| No | 79 (67.5%) | 35 (29.9%) | 44 (37.6%) | 3.649 |
| Yes | 38 (32.5%) | 24 (20.5%) | 14 (12.0%) | (0.056) |
| **HPV DNA** |  |  |  |  |
| Negative | 82 (70.1%) | 49 (41.9%) | 33 (28.2%) | **9.543** |
| Positive | 35 (29.9%) | 10 (8.5%) | 25 (21.4%) | **(0.002)** |
| **HPV mRNA ISH** |  |  |  |  |
| Negative | 87 (74.4%) | 51 (43.6%) | 36 (30.8%) | **9.112** |
| Positive | 30 (25.6%) | 8 (6.8%) | 22 (18.8%) | **(0.003)** |
| **p53/p16** |  |  |  |  |
| p53+/p16- | 54 (46.2%) | 34 (29.1%) | 20 (17.1%) | **7.891** |
| p53-/p16+ | 34 (29.1%) | 11 (9.4%) | 23 (19.7%) | **(0.019)** |
| p53-/p16- | 29 (24.8%) | 14 (12.0%) | 15 (12.8%) |  |
| **Recurrence of cancer** |  |  |  |  |
| No | 79 (67.5%) | 35 (29.9%) | 44 (37.6%) | 3.649 |
| Yes | 38 (32.5%) | 24 (20.5%) | 14 (12.0%) | (0.056) |
| **Death caused by VSCC** |  |  |  |  |
| No | 94 (80.3%) | 46 (39.3%) | 48 (41.0%) | 0.425 |
| Yes | 23 (19.7%) | 13 (11.1%) | 10 (8.5%) | (0.514) |

### Construction of immunoscore models with independent prognostic value for PFS and DSS

To further evaluate prognostic epithelial and stromal immune features and establish a combined immunoscore, the LASSO Cox regression model was used in which tuning parameter (λ) values were selected by 10-fold cross-validation (via minimum criteria) (Figure 3A, Supplementary Figure 5A). This identified four non-zero coefficients based on PFS and five non-zero coefficients based on DSS (Figure 3B, Supplementary Figure 5B). Immunoscores were subsequently calculated using these coefficients: immunoscore^PFS^ = (1.8609 x number of epithelial PD-L1+ cells/mm^2^) + (-4.5255 x number of epithelial CD8+ cells/mm^2^) + (2.1746 x number of epithelial FoxP3+ cells/mm^2^) + (0.1497 x number of stromal CD14+cells/mm^2^) x 10^-4^) and immunoscore^DSS^ = (0.2744 x number of stromal PD-L1+ cells/mm^2^) + (-0.1481 x number of epithelial CD8+ cells/mm^2^) + (-3.160 x number of epithelial FoxP3+ cells/mm^2^) + (-0.0143 x number of epithelial CD14+cells/mm^2^) + (0.1436 x number of stromal CD14+cells/mm^2^) x 10^-3^). Next, the association between immunoscores and survival outcomes was investigated. In multivariate Cox analysis, immunoscore^PFS^ and immunoscore^DSS^ were independent prognosticators of PFS (HR = 5.09, p = 0.001) and DSS (HR = 5.98, p = 0.007) respectively after adjusting for confounders (Figure 3C-D). When dichotomizing immunoscores using the maximally selected rank statistics method Kaplan Meier analysis showed that a high immunoscore^PFS^ was associated with poor PFS (log rank test: p < 0.001) and a high immunoscore^DSS^ predicted poor DSS (log rank test: p < 0.001) (Figure 3E). The VSCC cohort was then stratified by HPV mRNA ISH status to investigate the prognostic contribution of immunoscores within the HPV-independent subgroup. This analysis revealed that a low immunoscore^PFS^ and a low immunoscore^DSS^ significantly predicted favorable PFS (log rank test: p < 0.001) and DSS (log rank test: p < 0.001), respectively in HPV-independent tumors (Figure 3F). In contrast to the maximally selected rank statistics method, only immunoscore^DSS^ was significantly associated with DSS in the entire cohort (log rank test: p = 0.002) when immunoscores were dichotomized by median values (Figure 3G).

**Figure 3:**
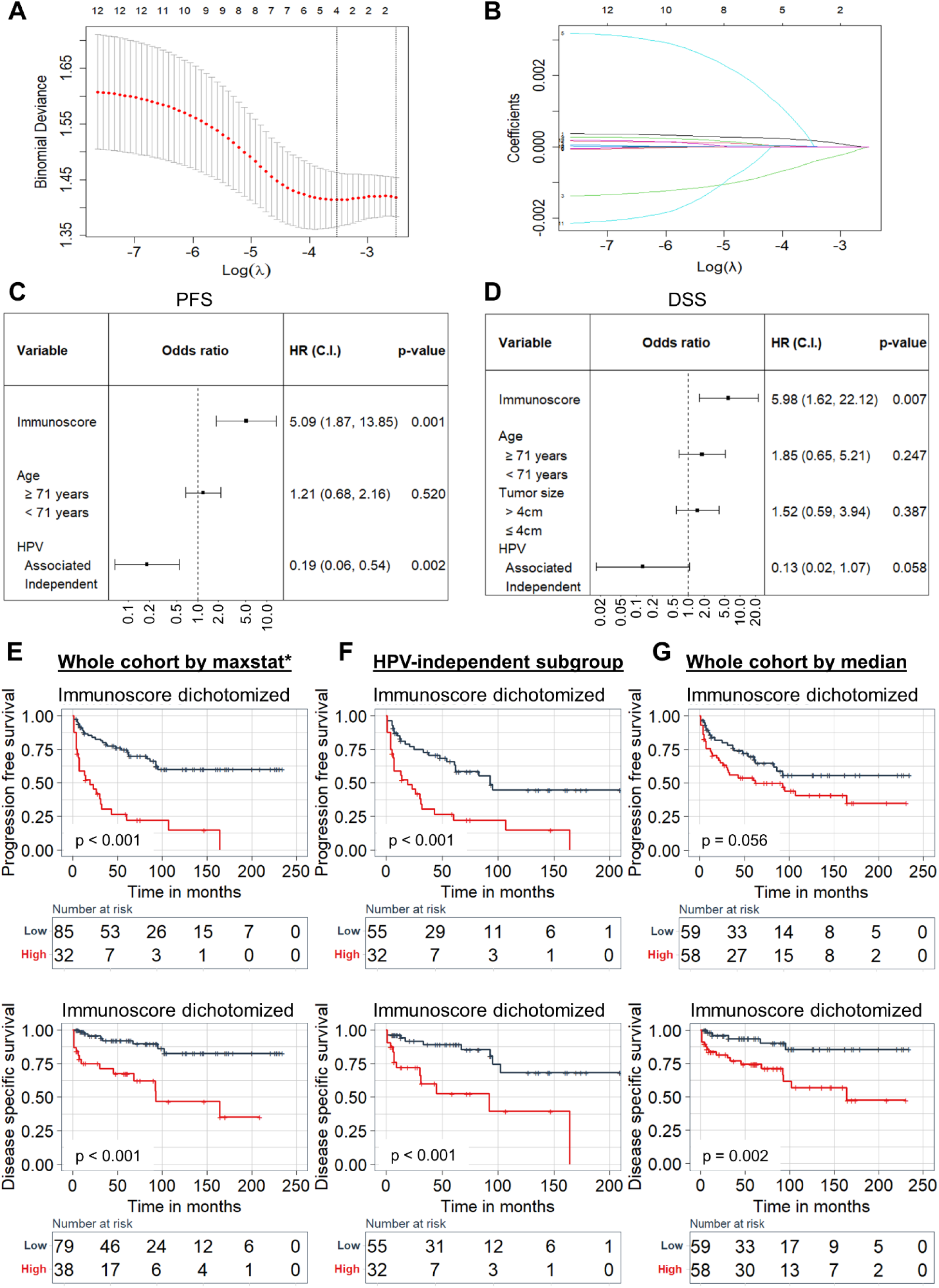
Determining immunoscores based on the LASSO cox regression model. (A) Immune feature selection for prediction of PFS by applying 10-fold cross-validation and training a LASSO regression model. The dotted vertical lines indicate selected λ values using minimum criteria. (B) Regression coefficient profiles of epithelial and stromal CD4+, CD8+, CD68+, CD14+, FoxP3+, and PD-L1+ subsets in the LASSO regression model. Forest plots demonstrating the association of immunescores and clinicopathological features on (D) PFS and (E) DSS. Hazard ratios (HR) with 95% confidence interval (C.I.) and p-values are shown. Kaplan-Meier curves showing the influence of immunoscores, dichotomized by the maximally selected rank statistics (maxstat*) method, on PFS and DSS for the (E) entire cohort and (F) HPV-independent subgroup. (G) Kaplan-Meier curves showing the influence of immunoscores, dichotomized by median values, on PFS and DSS for the entire cohort

### Integration of immunoscores with HPV status and validation of the prognostic models

Drawing upon the prognostic implications of immunoscores and HPV status, we amalgamated these variables to formulate nomograms termed as immunoscore-HPV (IS-HPV) index^PFS^ and IS-HPV index^DSS^ (Figure 4A). Using the nomograms, the probability of an outcome can be predicted by calculating a total score based on the points contributed by immunoscore and HPV status predictors. A bootstrap resampling approach was employed to generate 1,000 iterations for internal validation of the immunoscore and IS-HPV index models. Calibration plots indicated satisfactory agreement between observed outcomes and predicted outcomes from the nomograms. The IS-HPV index^PFS^ and IS-HPV index^DSS^ concordance (c)-index were 0.752 (corrected c-index: 0.750) and 0.760 (corrected c-index: 0.752) for PFS and DSS, respectively (Figure 4B). Comparatively, the corrected c-index for HPV status-based models alone was 0.666 for PFS and 0.631 for DSS. For immunoscore^PFS^ and immunoscore^DSS^ based models the corrected c-index were 0.667 and 0.715 for PFS and DSS respectively. To further assess the efficacy of the nomograms an IS-HPV index score was computed for all the patients in the cohort based on coefficients derived from logistic regression models used to construct the nomograms: IS-HPV index^PFS^ score = (1.558 x immunoscore) + (-1.931 x HPV status) and IS-HPV index^DSS^ score = (2.152 x immunoscore) + (- 2.066 x HPV status). The IS-HPV index scores were then dichotomized based on the maximally selected rank statistics method to select a cutoff (-0.430 for IS-HPV index^PFS^ and -0.028 for IS-HPV index^DSS^) for Kaplan-Meier analysis. High IS-HPV index^PFS^ and IS-HPV index^DSS^ scores were associated with poor PFS (log rank test: p < 0.001) and DSS (log rank test: p < 0.001) respectively (Figure 4C) and an improved discriminatory ability was observed for DSS when comparing the IS-HPV index^DSS^ to HPV status alone (Supplementary Figure 6). Furthermore, decision curve analysis demonstrated a clear net benefit from using the IS-HPV index^PFS^ nomogram when the threshold probability exceeded 0.15 PFS, and consistently across all threshold probabilities for DSS. At clinically relevant threshold probabilities for DSS, the integrated IS-HPV index^DSS^ model surpassed the performance of the immunoscore and HPV status models individually (Figure 4D).

**Figure 4:**
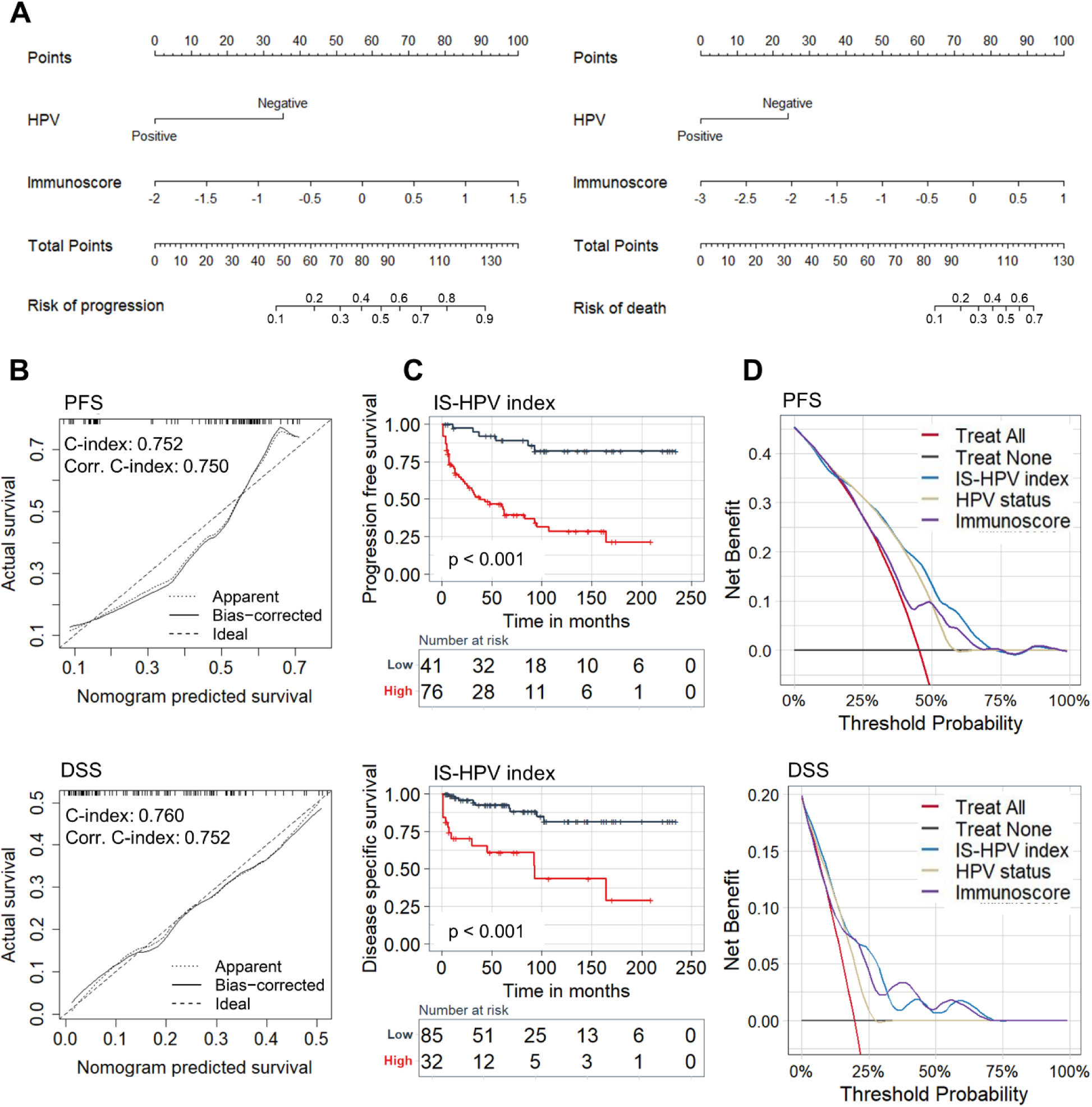
Construction of immunoscore-HPV index nomograms. (A) Immunoscore-HPV (IS-HPV) index nomograms for PFS and DSS. (B) Calibration plots demonstrating the agreement between nomogram predicted outcomes and actual survival outcomes. Concordance index (c-index) and corrected (corr) c-index values are provided. IS-HPV index scores were calculated for the entire VSCC cohort using coefficients from the logistic regression model and dichotomized based on the maximally selected rank statistics method. (C) Kaplan-Meier curves showing the influence of IS-HPV index on PFS and DSS. (D) Decision curve analysis for the nomograms for PFS and DSS.

## Discussion

It is becoming increasingly evident that the efficacy of immunotherapeutic modalities in cancer is affected by the ability to establish an enduring immune response. Indeed, the local immune infiltrate undergoes dynamic changes following treatment and may be indicative of prognosis in cancer (12). The association between *in-situ* infiltration of immune cell subsets and survival in VSCC remains unclear and the search for novel biomarkers continues. Here, we developed the first immunoscore model for VSCC by interrogating PD-L1 expression and immune cell infiltration patterns in the epithelium and stroma of tumors. Our analysis revealed that immunoscores independently predict survival in VSCC providing superior prognostication compared to immunological features assessed alone. Furthermore, integrating immunoscores with HPV status and establishing an IS-HPV index improved the prognostic impact of the models.

VSCC manifests through two distinct molecular pathways: HPV-independent and HPV-associated disease (25). Studies have shown that HPV-independent tumors are associated with poorer survival (10, 11, 26) and novel biomarkers are needed to categorize these cases. Immunoscores are prognostic tools focused on tumor immune interactions, that may contribute additional clinical value to traditional staging systems (27). Even though combined risk factor models have been presented for VSCC (28, 29), immunoscores have yet to be explored. In this study low immunoscores corresponded with improved PFS and DSS respectively in HPV-independent tumors suggesting that clinical use of immunoscores could improve risk stratification in VSCC. Moreover, immunoscores were independent predictors of survival in the entire VSCC cohort, corroborating findings from other cancer types (30, 31), although the immunological features used to determine the models differed. Previously, immunoscores were established based on infiltration of T cells (19, 27, 32) however in this study myeloid subsets were also included to more accurately reflect the immune landscape. Besides the number of infiltrating immune cells, studies have also shown that the spatiotemporal dynamics influence disease progression (33). We therefore spatially resolved immunological features and investigated the epithelial or stromal specific contribution instead of only focusing on the density of cells (34). To accomplish this, IHC and digital quantification were used to determine immunoscores. The immunoscores were then integrated with HPV status to form easy-to-use nomograms. At clinically relevant threshold probabilities for DSS, our data indicates that using the nomogram may offer additional clinical benefit to VSCC patients compared to HPV status or immunoscore based models alone. Similarly, He et al demonstrated that combining immunoscores with clinicopathological features improved the impact of the models in non-small cell lung carcinoma (NSCLC) (35). Taken together, our findings suggest that the IS-HPV index model could be a rapid and cost-effective diagnostic test to improve risk stratification in VSCC and may enable selection of patients who are likely to benefit from less aggressive treatment regimens. Furthermore, the immunoscores may demonstrate predictive value for immunotherapy, as recently reported for NSCLC (36), however we were unable to assess this in our cohort as patients had not received immunotherapy.

We also assessed the immunological features used to determine immunoscores individually. PD-L1 is pivotal in cancer immunoediting and the expression level impacts patient survival outcomes in many cancer types (37). Consistent with Hecking et al, our data indicates that positive PD-L1 status (TPS > 1) is an independent prognostic factor associated with poor survival, HPV-independent disease (17), and the p53+/p16-subtype. Other studies found no association between epithelial PD-L1 expression and survival (38, 39). The discrepancies may stem from variations in sample size or methodologies and thresholds used to define PD-L1 status. Considering the rarity of this disease, a relatively large cohort was recruited in this study and PD-L1 expression was determined by IHC and digital image analysis instead of manual scoring or genomic profiling. PD-L1 status was determined by TPS > 1 contrasting other studies where higher thresholds were used (18, 39, 40). Future work is needed to harmonize scoring and cut-offs to evaluate the prognostic importance of PD-L1 in VSCC. Notably, PD-L1 status was discordant when comparing primary tumors to corresponding recurrent or metastatic lesions, aligning with previous reports (38), indicating it may be important to retest this marker in metastatic or recurrent VSCC.

Our data revealed a strong correlation between the number of PD-L1+ cells and infiltration of CD68+ macrophages in both the epithelium and stroma of VSCC. Studies have suggested that tumor associated macrophages (TAMs) are the predominant immune cells that express PD-L1 along with cancer cells in NSCLC and breast cancer (41, 42). The strong correlation we observed may therefore be due to the presence of TAMs expressing CD68+ and PD-L1+ surface markers. Additionally, high stromal infiltration of CD68+ macrophages was linked to HPV-independent tumors and poorer survival outcomes. Condic et al. reported that CD163+ TAMs were an unfavorable prognostic variable in VSCC (43). While CD68+ serves as a universal marker for macrophages, CD163+ is predominantly expressed by the M2-polarized subtype. Similar to CD68+ macrophages, high stromal CD14+ monocyte infiltration was linked to poor survival and HPV-independent pathogenesis. Van Esch et al showed no significant association between stromal CD14+ infiltration and survival in vulvar epithelial neoplasia lesions, however HPV-independent VSCC cases which constitute most of our cohort, were not assessed in their study (44). In contrast, Abdulrahman et al reported that a strong epithelial CD14+ infiltration was associated with improved survival (45). Importantly, these cells were negative for other myeloid markers while our study consisted of a mixed population of CD14+ positive cells. Future work should aim to decipher the macrophage and monocyte subtypes in our cohort.

Lymphocytes, particularly CD8+ T cells, are essential effectors in the adaptive immune response against tumors (46). In accordance with other studies, we found no correlation between epithelial CD8+ T cell infiltration and survival in VSCC (47). Conversely, Kortekaas, et al. reported that CD3+CD8+Foxp3− T cell infiltration in the epithelial compartments was correlated with favorable survival (15). An earlier study reported no association between this T cell subset and survival, irrespective of dichotomization using median values or ROC analysis (16) highlighting the need for further research. Inconsistent with our results, the number of CD3+CD8+FoxP3- and CD3+CD8-FoxP3+ T cells were significantly different when comparing HPV-independent and HPV-associated disease in the abovementioned study. The discrepancies may be due to differences in the investigated immune cell subtypes. Although epithelial infiltration of FoxP3+ T cells was associated with improved DSS, independent prognostic value was not observed in this study conflicting with another report (48). Here, the prognostic impact of FoxP3+ was adjusted for HPV mRNA ISH instead of p16 IHC as a surrogate marker for HPV. Our group recently demonstrated differences concerning HPV mRNA ISH and p16 IHC for prediction of survival in VSCC (10).

This study has limitations such as the retrospective study design and the recruitment of all participants from one single center. To mitigate the limitations, thorough internal validation was conducted. However, multicenter prospective studies are necessary to further confirm the generalizability of our findings. In this study PD-L1 expression and immune cell infiltration were investigated using IHC and digital image analysis based on established algorithms. A key advantage of this approach is the acquisition of data through unbiased measurements. However, the method faces challenges, including difficulties applying automated workflows to produce accurate results due to biomarker heterogeneity and divergences in tissue samples. Routine calibration with a pathologist was performed to ensure analysis validity. Furthermore, data acquired from TMA cores may not be biologically representative due to intra-tumor heterogeneity. We attempted to mitigate the impact of this by averaging two or three cores per case for downstream analysis. Additionally, our data demonstrated that the method of dichotomization may influence analysis outcomes. Future studies are therefore required to determine the optimal approach for dichotomizing biomarkers to binary variables (49).

In summary, this study provides a comprehensive analysis of PD-L1 expression and immune cell infiltration in epithelial and stromal compartments of VSCC. For the first time, we introduce a VSCC immunoscore designed to enhance disease risk stratification. By integrating this immunoscore with HPV status, determined via HPV mRNA ISH, we established an immunoscore-HPV index for prediction of PFS and DSS in VSCC. Additional research is necessary to confirm the accuracy and clinical utility of the immunoscore-HPV index models.

## Data Availability

Data associated with the findings of this study are included in the manuscript or in the uploaded supplementary files and are available from the corresponding author upon reasonable request.

## Abbreviations

C-INDEX: concordance-index
DSS: disease specific survival
FFPE: formalin-fixed paraffin embedded
FIGO: the International Federation of Gynecology and Obstetrics
HPV: human papilloma virus
HUS: Haukeland University Hospital
IHC: immunohistochemistry
ISH: in situ hybridization
IS-HPV: immunoscore-human papilloma virus index
LASSO: least absolute shrinkage and selection operator
PCR: polymerase chain reaction
PD-1: programmed cell death protein-1
PD-L1: programmed death ligand protein-1
PFS: progression free survival
REK: Regional Committee for Medical and Health Research Ethics
TIME: tumor immune microenvironment
TMA: tissue microarray
VSCC: vulva squamous cell carcinoma

## Declarations

### Ethics approval and consent to participate

This study was approved by the Regional Committee for Medical and Health Research Ethics Norway (REK Nos: 2017/279 and 2014/1907). Written consent was obtained from all participants included in this study.

### Consent for publication

Not applicable

### Availability of data and materials

Data associated with the findings of this study are included in the article or in the uploaded supplementary files and are available from the corresponding author upon reasonable request.

### Authors’ contributions

Conceptualization, RE, DEC, LB and HND; data curation, RE, LB and HND; formal analysis, RE and HND; funding acquisition, HND, LB and DEC; investigation, RE, IHH, MF, IBMK, LCVT and HND; methodology, RE, IHH, MF and HND; project administration, DEC, LB and HND; supervision, DEC, LB and HND; validation, RE and HND; visualization, RE, IBMK, LVCT; writing—original draft, RE and HND; writing—review & editing, RE, HND, DEC and LB. All authors have read and agreed to the published version of the manuscript.

## Acknowledgements

We would like to thank all the patients who participated in this study.

## References

1. Bray F, Laversanne M, Sung H, Ferlay J, Siegel RL, Soerjomataram I, et al. Global cancer statistics 2022: GLOBOCAN estimates of incidence and mortality worldwide for 36 cancers in 185 countries. CA Cancer J Clin. 2024;74(3):229-63.

2. Siegel RL, Giaquinto AN, Jemal A. Cancer statistics, 2024. CA Cancer J Clin. 2024;74(1):12-49.

3. Meltzer-Gunnes CJ, Lie AK, Jonassen CGM, Rangberg A, Nystrand CF, Smastuen MC, et al. Time trends in human papillomavirus prevalence and genotype distribution in vulvar carcinoma in Norway. Acta Obstet Gynecol Scand. 2024;103(1):153–64.

4. Zach D, Avall-Lundqvist E, Falconer H, Hellman K, Johansson H, Floter Radestad A. Patterns of recurrence and survival in vulvar cancer: A nationwide population-based study. Gynecol Oncol. 2021;161(3):748–54.

5. Nooij LS, Brand FA, Gaarenstroom KN, Creutzberg CL, de Hullu JA, van Poelgeest MI. Risk factors and treatment for recurrent vulvar squamous cell carcinoma. Crit Rev Oncol Hematol. 2016;106:1–13.

6. Schnack TH, Froeding LP, Kristensen E, Niemann I, Ortoft G, Hogdall E, et al. Preoperative predictors of inguinal lymph node metastases in vulvar cancer - A nationwide study. Gynecol Oncol. 2022;165(3):420–7.

7. Soderini A, Aragona A, Reed N. Advanced Vulvar Cancers: What are the Best Options for Treatment? Curr Oncol Rep. 2016;18(10):64.

8. Weberpals JI, Lo B, Duciaume MM, Spaans JN, Clancy AA, Dimitroulakos J, et al. Vulvar Squamous Cell Carcinoma (VSCC) as Two Diseases: HPV Status Identifies Distinct Mutational Profiles Including Oncogenic Fibroblast Growth Factor Receptor 3. Clin Cancer Res. 2017;23(15):4501–10.

9. Hinten F, Molijn A, Eckhardt L, Massuger L, Quint W, Bult P, et al. Vulvar cancer: Two pathways with different localization and prognosis. Gynecol Oncol. 2018;149(2):310–7.

10. Dongre HN, Elnour R, Tornaas S, Fromreide S, Thomsen LCV, Kolseth IBM, et al. TP53 mutation and human papilloma virus status as independent prognostic factors in a Norwegian cohort of vulva squamous cell carcinoma. Acta Obstet Gynecol Scand. 2024;103(1):165–75.

11. Kortekaas KE, Bastiaannet E, van Doorn HC, de Vos van Steenwijk PJ, Ewing-Graham PC, Creutzberg CL, et al. Vulvar cancer subclassification by HPV and p53 status results in three clinically distinct subtypes. Gynecol Oncol. 2020;159(3):649–56.

12. Bruni D, Angell HK, Galon J. The immune contexture and Immunoscore in cancer prognosis and therapeutic efficacy. Nat Rev Cancer. 2020;20(11):662–80.

13. Santoro A, Angelico G, Inzani F, Arciuolo D, d’Amati A, Addante F, et al. The emerging and challenging role of PD-L1 in patients with gynecological cancers: An updating review with clinico-pathological considerations. Gynecol Oncol. 2024;184:57-66.

14. Schwab R, Schiestl LJ, Cascant Ortolano L, Klecker PH, Schmidt MW, Almstedt K, et al. Efficacy of pembrolizumab in advanced cancer of the vulva: a systematic review and single-arm meta-analysis. Front Oncol. 2024;14:1352975.

15. Kortekaas KE, Santegoets SJ, Tas L, Ehsan I, Charoentong P, van Doorn HC, et al. Primary vulvar squamous cell carcinomas with high T cell infiltration and active immune signaling are potential candidates for neoadjuvant PD-1/PD-L1 immunotherapy. J Immunother Cancer. 2021;9(10).

16. Kortekaas KE, Santegoets SJ, Abdulrahman Z, van Ham VJ, van der Tol M, Ehsan I, et al. High numbers of activated helper T cells are associated with better clinical outcome in early stage vulvar cancer, irrespective of HPV or p53 status. J Immunother Cancer. 2019;7(1):236.

17. Hecking T, Thiesler T, Schiller C, Lunkenheimer JM, Ayub TH, Rohr A, et al. Tumoral PD-L1 expression defines a subgroup of poor-prognosis vulvar carcinomas with non-viral etiology. Oncotarget. 2017;8(54):92890–903.

18. Sznurkowski JJ, Zawrocki A, Sznurkowska K, Peksa R, Biernat W. PD-L1 expression on immune cells is a favorable prognostic factor for vulvar squamous cell carcinoma patients. Oncotarget. 2017;8(52):89903–12.

19. Pages F, Mlecnik B, Marliot F, Bindea G, Ou FS, Bifulco C, et al. International validation of the consensus Immunoscore for the classification of colon cancer: a prognostic and accuracy study. Lancet. 2018;391(10135):2128–39.

20. Pages F, Andre T, Taieb J, Vernerey D, Henriques J, Borg C, et al. Prognostic and predictive value of the Immunoscore in stage III colon cancer patients treated with oxaliplatin in the prospective IDEA France PRODIGE-GERCOR cohort study. Ann Oncol. 2020;31(7):921–9.

21. Argiles G, Tabernero J, Labianca R, Hochhauser D, Salazar R, Iveson T, et al. Localised colon cancer: ESMO Clinical Practice Guidelines for diagnosis, treatment and follow-up. Ann Oncol. 2020;31(10):1291–305.

22. Haave H, Gulati S, Brekke J, Lybak S, Vintermyr OK, Aarstad HJ. Tumor stromal desmoplasia and inflammatory response uniquely predict survival with and without stratification for HPV tumor infection in OPSCC patients. Acta Otolaryngol. 2018;138(11):1035–42.

23. Bankhead P, Loughrey MB, Fernandez JA, Dombrowski Y, McArt DG, Dunne PD, et al. QuPath: Open source software for digital pathology image analysis. Sci Rep. 2017;7(1):16878.

24. Davis AA, Patel VG. The role of PD-L1 expression as a predictive biomarker: an analysis of all US Food and Drug Administration (FDA) approvals of immune checkpoint inhibitors. J Immunother Cancer. 2019;7(1):278.

25. Clancy AA, Spaans JN, Weberpals JI. The forgotten woman’s cancer: vulvar squamous cell carcinoma (VSCC) and a targeted approach to therapy. Ann Oncol. 2016;27(9):1696–705.

26. Allo G, Yap ML, Cuartero J, Milosevic M, Ferguson S, Mackay H, et al. HPV-independent Vulvar Squamous Cell Carcinoma is Associated With Significantly Worse Prognosis Compared With HPV-associated Tumors. Int J Gynecol Pathol. 2020;39(4):391–9.

27. Angell HK, Bruni D, Barrett JC, Herbst R, Galon J. The Immunoscore: Colon Cancer and Beyond. Clin Cancer Res. 2020;26(2):332–9.

28. Zhang T, Zhu Y, Luo J, Li J, Niu S, Chen H, et al. An integrated model for prognosis in vulvar squamous cell carcinoma. BMC Cancer. 2023;23(1):534.

29. Zhao Z, Zhen S, Liu N, Ding D, Zhang D, Kong J. Survival nomograms for vulvar squamous cell carcinoma based on the SEER database and a Chinese external validation cohort. Int J Gynaecol Obstet. 2024;165(3):1130–43.

30. Ghiringhelli F, Bibeau F, Greillier L, Fumet JD, Ilie A, Monville F, et al. Immunoscore immune checkpoint using spatial quantitative analysis of CD8 and PD-L1 markers is predictive of the efficacy of anti-PD1/PD-L1 immunotherapy in non-small cell lung cancer. EBioMedicine. 2023;92:104633.

31. Mezheyeuski A, Backman M, Mattsson J, Martin-Bernabe A, Larsson C, Hrynchyk I, et al. An immune score reflecting pro- and anti-tumoural balance of tumour microenvironment has major prognostic impact and predicts immunotherapy response in solid cancers. EBioMedicine. 2023;88:104452.

32. Han B, Yim J, Lim S, Na S, Lee C, Kim TM, et al. Prognostic Impact of the Immunoscore Based on Whole-Slide Image Analysis of CD3+ Tumor-Infiltrating Lymphocytes in Diffuse Large B-Cell Lymphoma. Mod Pathol. 2023;36(9):100224.

33. Mezheyeuski A, Bergsland CH, Backman M, Djureinovic D, Sjoblom T, Bruun J, et al. Multispectral imaging for quantitative and compartment-specific immune infiltrates reveals distinct immune profiles that classify lung cancer patients. J Pathol. 2018;244(4):421–31.

34. Nie RC, Yuan SQ, Wang Y, Chen YB, Cai YY, Chen S, et al. Robust immunoscore model to predict the response to anti-PD1 therapy in melanoma. Aging (Albany NY). 2019;11(23):11576–90.

35. He L, Huang Y, Chen X, Huang X, Wang H, Zhang Y, et al. Development and Validation of an Immune-Based Prognostic Risk Score for Patients With Resected Non-Small Cell Lung Cancer. Front Immunol. 2022;13:835630.

36. Hijazi A, Antoniotti C, Cremolini C, Galon J. Light on life: immunoscore immune-checkpoint, a predictor of immunotherapy response. Oncoimmunology. 2023;12(1):2243169.

37. Schreiber RD, Old LJ, Smyth MJ. Cancer immunoediting: integrating immunity’s roles in cancer suppression and promotion. Science. 2011;331(6024):1565-70.

38. Lerias S, Esteves S, Silva F, Cunha M, Cochicho D, Martins L, et al. CD274 (PD-L1), CDKN2A (p16), TP53, and EGFR immunohistochemical profile in primary, recurrent and metastatic vulvar cancer. Mod Pathol. 2020;33(5):893-904.

39. Thangarajah F, Morgenstern B, Pahmeyer C, Schiffmann LM, Puppe J, Mallmann P, et al. Clinical impact of PD-L1 and PD-1 expression in squamous cell cancer of the vulva. J Cancer Res Clin Oncol. 2019;145(6):1651–60.

40. Czogalla B, Pham D, Trillsch F, Rottmann M, Gallwas J, Burges A, et al. PD-L1 expression and survival in p16-negative and -positive squamous cell carcinomas of the vulva. J Cancer Res Clin Oncol. 2020;146(3):569–77.

41. Liu Y, Zugazagoitia J, Ahmed FS, Henick BS, Gettinger SN, Herbst RS, et al. Immune Cell PD-L1 Colocalizes with Macrophages and Is Associated with Outcome in PD-1 Pathway Blockade Therapy. Clin Cancer Res. 2020;26(4):970–7.

42. Wang L, Guo W, Guo Z, Yu J, Tan J, Simons DL, et al. PD-L1-expressing tumor-associated macrophages are immunostimulatory and associate with good clinical outcome in human breast cancer. Cell Rep Med. 2024;5(2):101420.

43. Condic M, Rohr A, Riemann S, Staerk C, Ayub TH, Doeser A, et al. Immune Profiling of Vulvar Squamous Cell Cancer Discovers a Macrophage-rich Subtype Associated with Poor Prognosis. Cancer Res Commun. 2024;4(3):861–75.

44. van Esch EM, van Poelgeest MI, Trimbos JB, Fleuren GJ, Jordanova ES, van der Burg SH. Intraepithelial macrophage infiltration is related to a high number of regulatory T cells and promotes a progressive course of HPV-induced vulvar neoplasia. Int J Cancer. 2015;136(4):E85–94.

45. Abdulrahman Z, Kortekaas KE, Welters MJP, van Poelgeest MIE, van der Burg SH. Monocyte infiltration is an independent positive prognostic biomarker in vulvar squamous cell carcinoma. Cancer Immunol Immunother. 2024;73(9):166.

46. Raskov H, Orhan A, Christensen JP, Gogenur I. Cytotoxic CD8(+) T cells in cancer and cancer immunotherapy. Br J Cancer. 2021;124(2):359–67.

47. Burandt E, Blessin NC, Rolschewski AC, Lutz F, Mandelkow T, Yang C, et al. T-Cell Density at the Invasive Margin and Immune Phenotypes Predict Outcome in Vulvar Squamous Cell Cancer. Cancers (Basel). 2022;14(17).

48. Arik D, Benli T, Telli E. Number of FoxP3+ regulatory T-cells are associated with recurrence in vulvar squamous cell carcinoma. J Gynecol Oncol. 2023;34(2):e16.

49. Prince Nelson SL, Ramakrishnan V, Nietert PJ, Kamen DL, Ramos PS, Wolf BJ. An evaluation of common methods for dichotomization of continuous variables to discriminate disease status. Commun Stat Theory Methods. 2017;46(21):10823–34.

