## Supplementary figures for "Prognostic value of an integrated human papilloma virus and immunoscore model to predict survival in vulva squamous cell carcinoma"

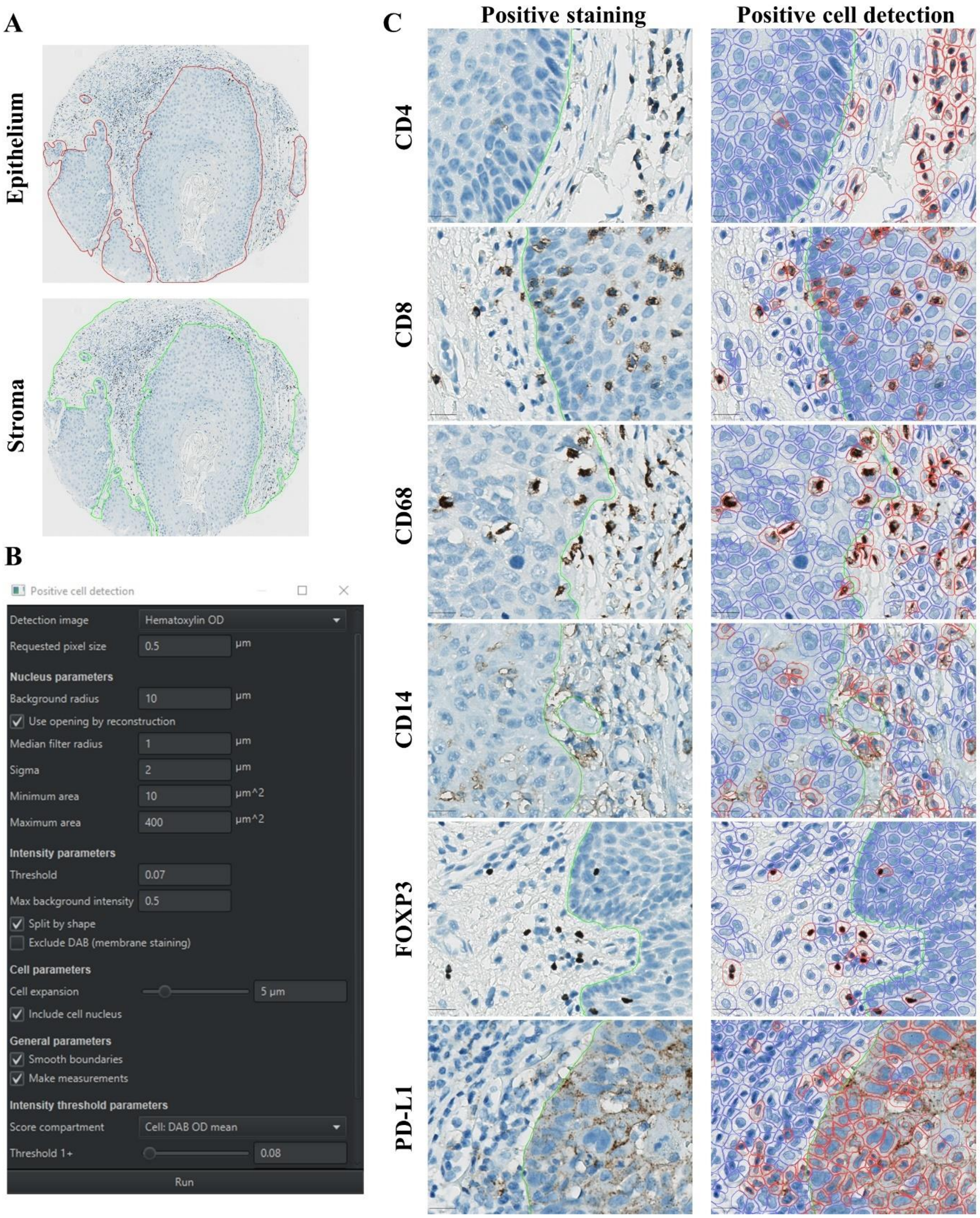


**Supplementary Figure 1:** QuPath analysis strategy. A) Annotation mark-up for epithelial and stromal compartments in TMA cores. B) QuPath input parameters for positive cell detection based on IHC and DAB staining. C) Depiction of DAB positive staining and corresponding positive cell detection for each marker.

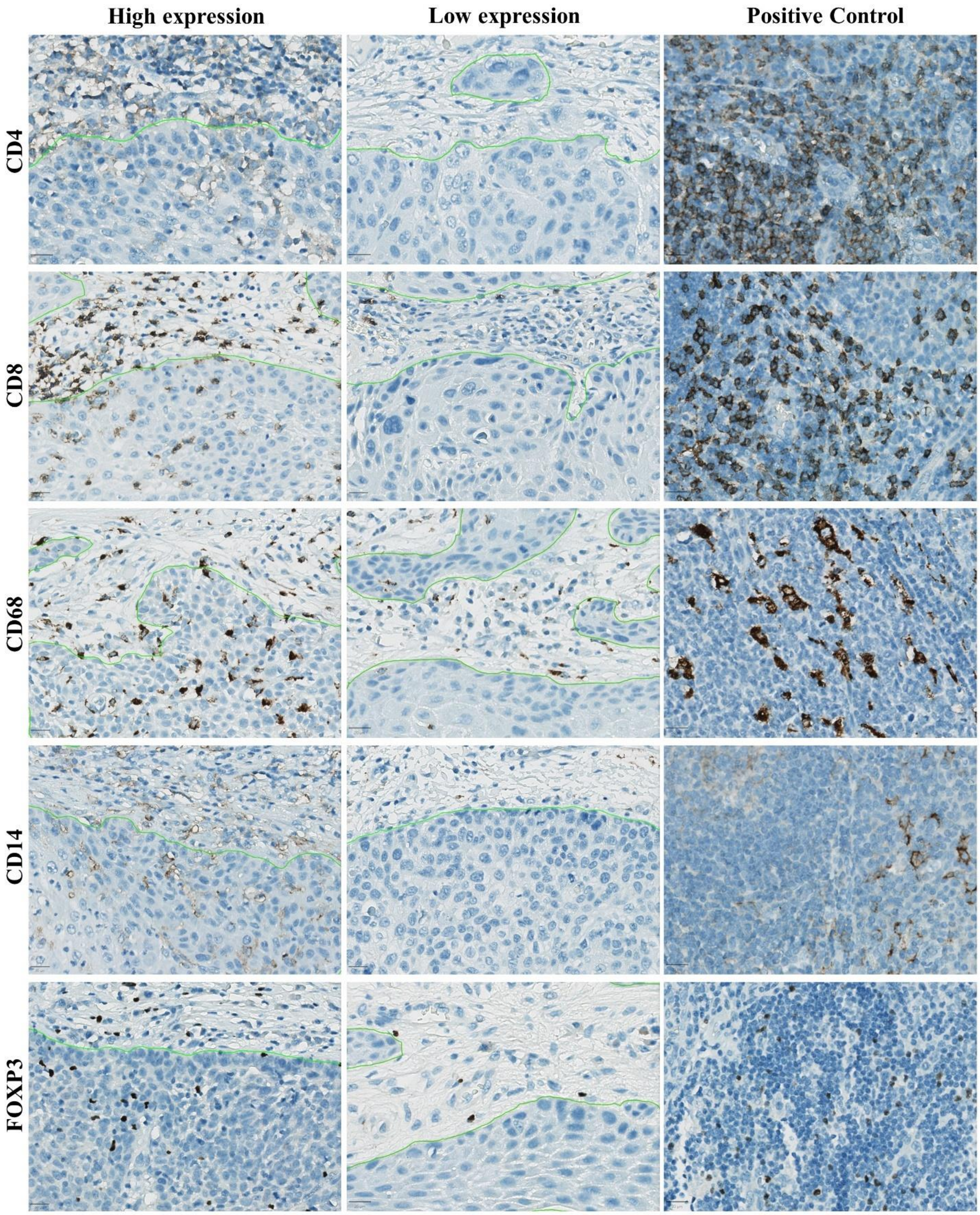


**Supplementary Figure 2:** Representative images of IHC staining demonstrating high and low expression of each immune cell subset. Subsets were stratified into high and low expression based on median values for each respective marker.

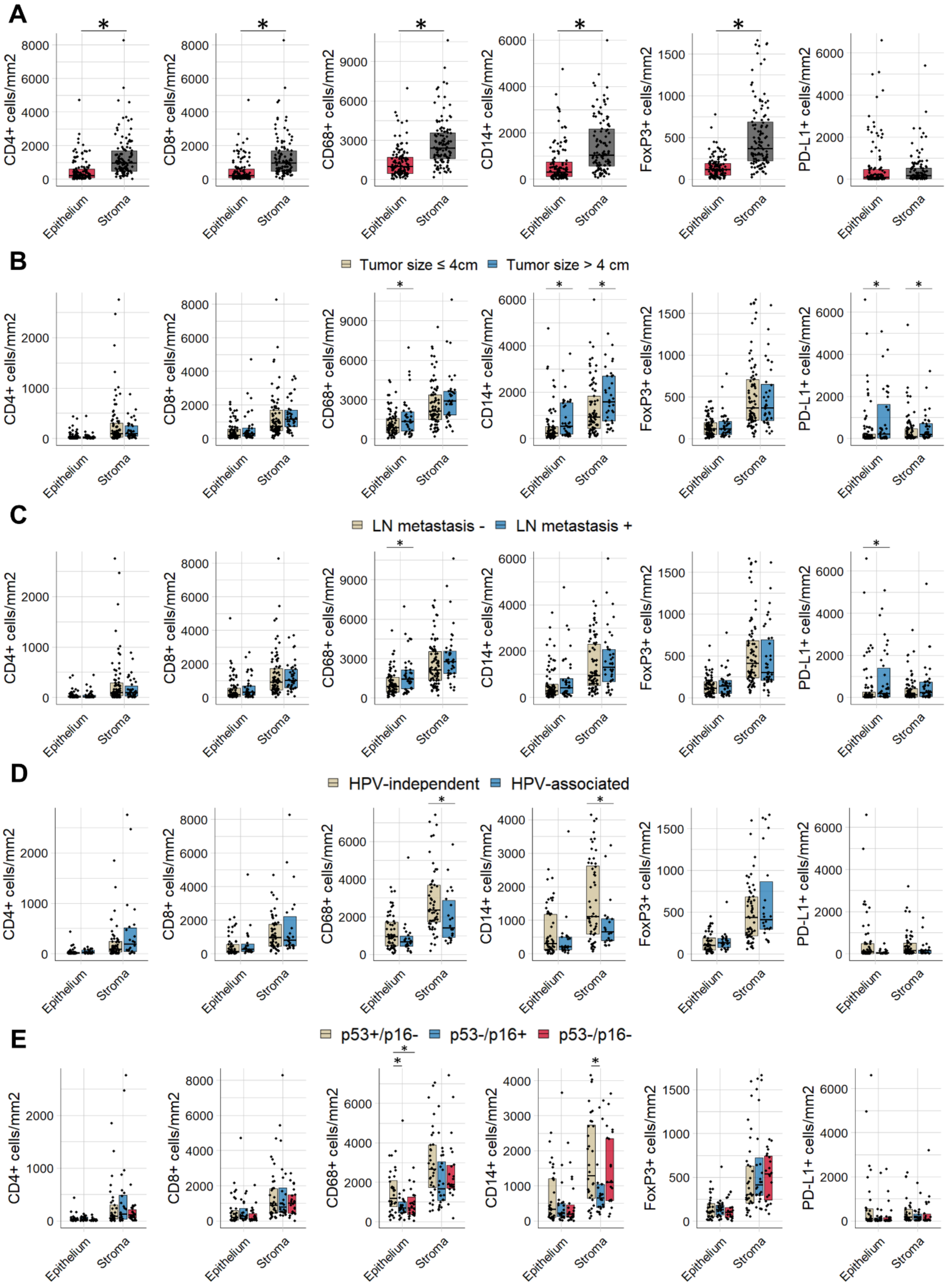
A
B
C
D
E
Tumor size ≤ 4cm
Tumor size > 4 cm
LN metastasis -
LN metastasis +
HPV-independent
HPV-associated
p53+/p16-
p53-/p16+
p53-/p16-
CD4+ cells/mm2
CD8+ cells/mm2
CD68+ cells/mm2
CD14+ cells/mm2
FoxP3+ cells/mm2
PD-L1+ cells/mm2
Epithelium
Stroma

**Supplementary Figure 3:** Box plots depicting differences in the number of cells per/mm$^2$ based on (A) epithelial and stromal compartments, (B) tumor size, and (C) lymph node metastasis in the entire cohort. Box plots depicting differences in the number of cells per/mm$^2$ based on (D) HPV status, as determined by HPV mRNA ISH and (E) p53/p16 subtypes in FIGO stage I/II VSCC. The bars indicate the interquartile range, and the median is highlighted with a black line. Individual cases are represented by black dots. p-values $< 0.05$ were considered statistically significant.

A

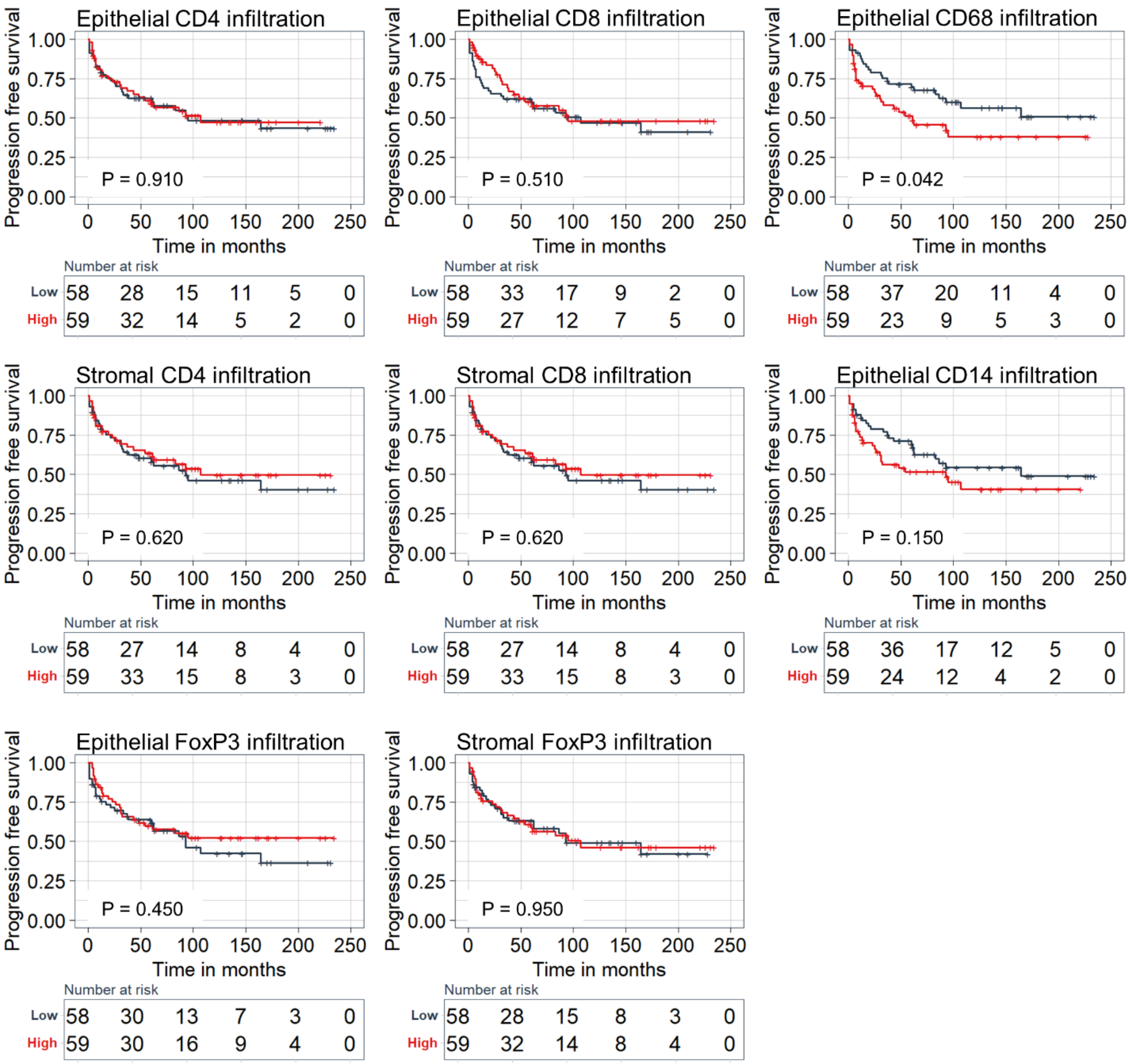
Epithelial CD4 infiltration
P = 0.910
Number at risk
Low 58 28 15 11 5 0
High 59 32 14 5 2 0
Epithelial CD8 infiltration
P = 0.510
Number at risk
Low 58 33 17 9 2 0
High 59 27 12 7 5 0
Epithelial CD68 infiltration
P = 0.042
Number at risk
Low 58 37 20 11 4 0
High 59 23 9 5 3 0
Stromal CD4 infiltration
P = 0.620
Number at risk
Low 58 27 14 8 4 0
High 59 33 15 8 3 0
Stromal CD8 infiltration
P = 0.620
Number at risk
Low 58 27 14 8 4 0
High 59 33 15 8 3 0
Epithelial CD14 infiltration
P = 0.150
Number at risk
Low 58 36 17 12 5 0
High 59 24 12 4 2 0
Epithelial FoxP3 infiltration
P = 0.450
Number at risk
Low 58 30 13 7 3 0
High 59 30 16 9 4 0
Stromal FoxP3 infiltration
P = 0.950
Number at risk
Low 58 28 15 8 3 0
High 59 32 14 8 4 0
Progression free survival
Time in months

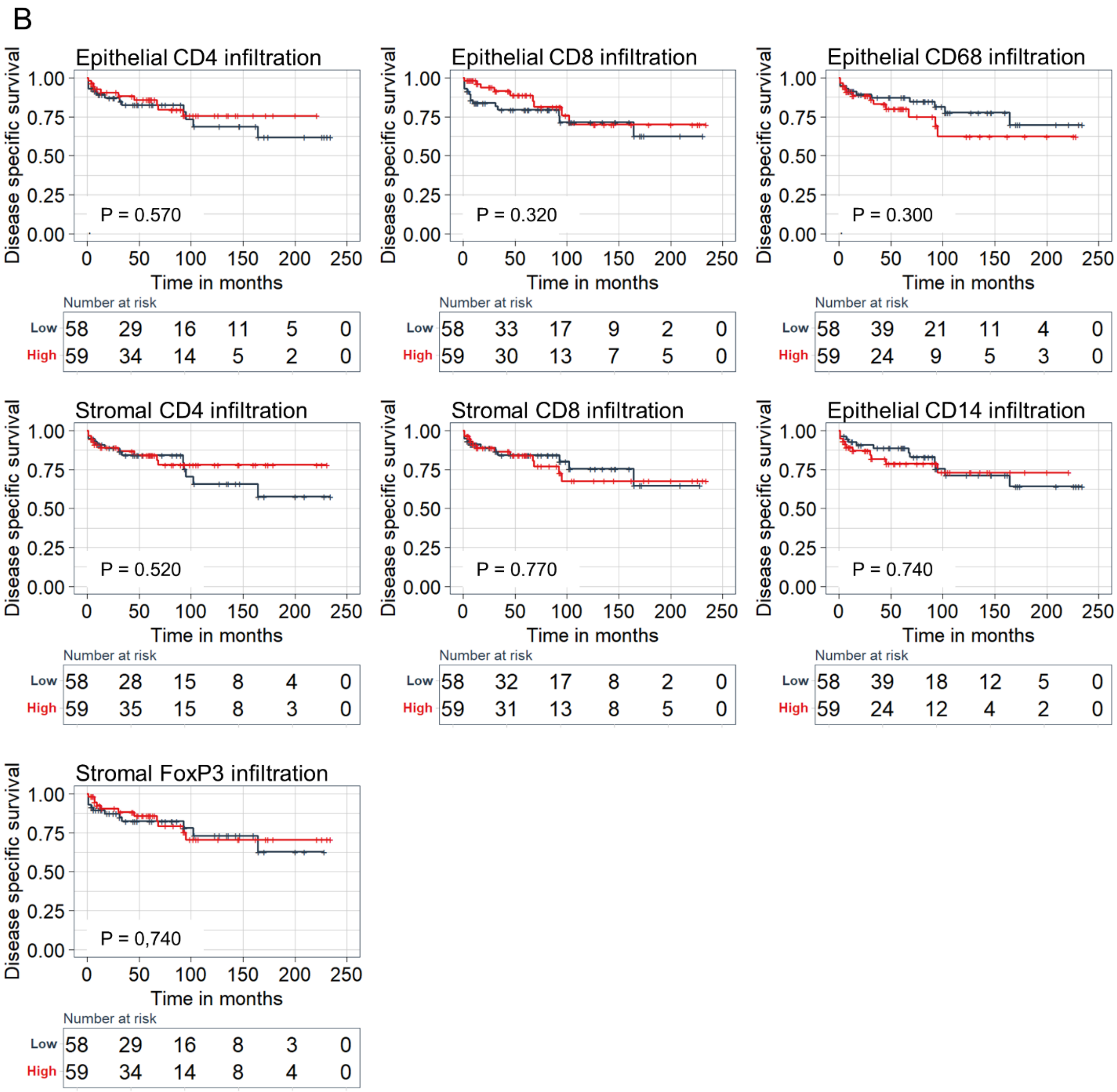


**Supplementary Figure 4:** Kaplan-Meier curves demonstrating influence of immune cell infiltration on (A) progression free- and (B) disease specific survival. p-values < 0.05 were considered statistically significant based on log rank test.

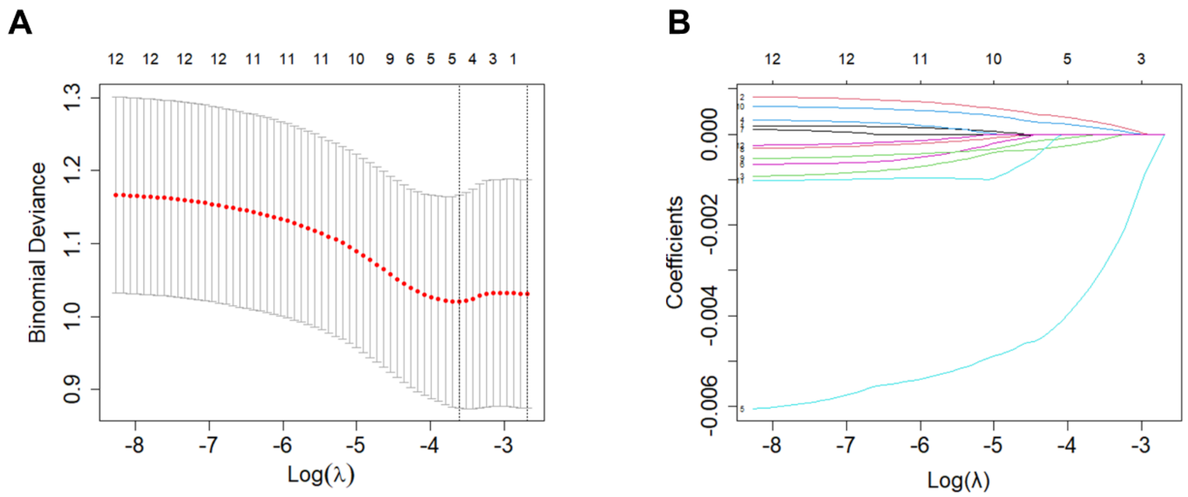


**Supplementary Figure 5:** Determining immunoscores based on the LASSO cox regression model. (A) Immune feature selection for prediction of DSS by applying 10-fold cross-validation and training a LASSO regression model. The dotted vertical lines indicate selected λ values using minimum criteria. (B) Regression coefficient profiles of epithelial and stromal CD4+, CD8+, CD68+, CD14+, FoxP3+, and PD-L1+ subsets in the LASSO regression model.

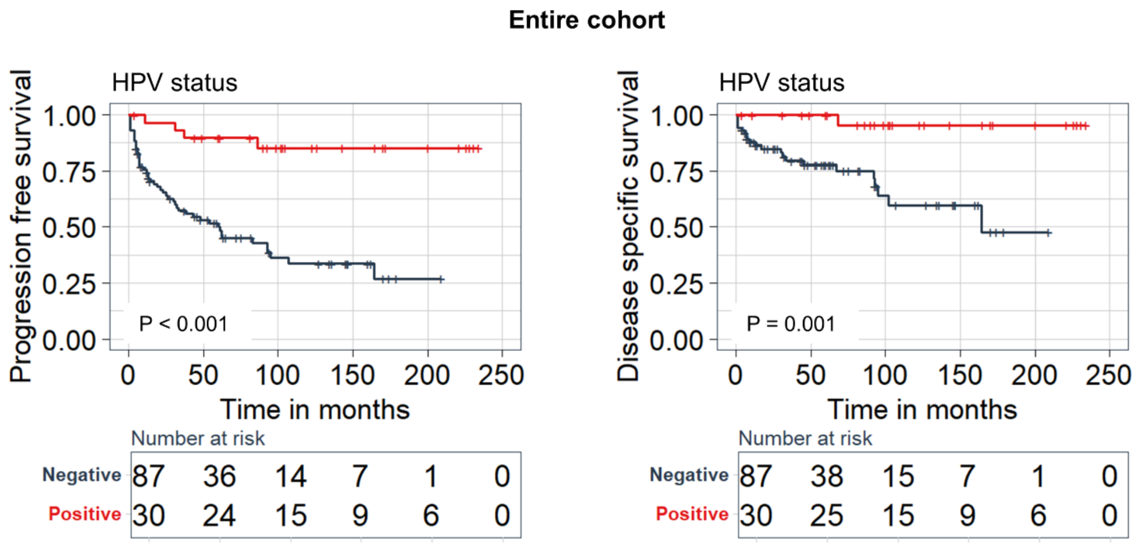


**Supplementary Figure 6:** Kaplan-Meier curves demonstrating influence of HPV status, as determined by HPV mRNA ISH, on progression free- and disease specific survival in the entire cohort. p-values < 0.05 were considered statistically significant based on log rank test.
