## Supplementary tables for "Prognostic value of an integrated human papilloma virus and immunoscore model to predict survival in vulva squamous cell carcinoma"

**Supplementary Table 1:** List of antibodies used for immunohistochemistry on the Ventana Benchmark ULTRA autostainer.

| Target | Clone | Antigen retrieval | Dilution | Incubation time | Incubation temperature | Source |
|---|---|---|---|---|---|---|
| CD4 | 1F6 | CC1 100°C, 48 min | 1:100 | 32 minutes | 36°C | Agilent (USA) |
| CD8 | C8/144B | CC1 95°C, 36 min | 1:100 | 32 minutes | 37°C | Agilent (USA) |
| CD68 | PG-M1 | CC1 95°C, 64 min | 1:100 | 32 minutes | 37°C | Agilent (USA) |
| CD14 | EPR3653 | CC1 100°C, 48 min | 1:100 | 32 minutes | 36°C | Cell Marque (USA) |
| FOXP3 | 259D/C7 | CC1 95°C, 64 min | 1:20 | 32 minutes | 37°C | BD Pharmingen (USA) |
| PD-L1 | 22C3 | CC1 100°C, 32 min | 1:50 | 32 minutes | 36°C | Dako/Agilent (USA) |

CC1: ULTRA Cell Conditioning Solution 1

**Supplementary Table 2:** Median values of immune cells and PD-L1+ cells in epithelial and stromal compartments of VSCC.

| Marker | Compartment | Median (number cells/mm$^2$) |
|---|---|---|
| **CD4** | Epithelium | 13.940 |
| | Stroma | 100.190 |
| **CD8** | Epithelium | 227.000 |
| | Stroma | 959.970 |
| **CD68** | Epithelium | 1002.255 |
| | Stroma | 2392.350 |
| **CD14** | Epithelium | 308.720 |
| | Stroma | 1040.620 |
| **FOXP3** | Epithelium | 116.520 |
| | Stroma | 367.550 |
| **PD-L1** | Epithelium | 76.440 |
| | Stroma | 162.955 |

**Supplementary Table 3:** Relationship between epithelial and stromal infiltration of immune cells with clinicopathological features.

| Clinicopathological features | CD4+ T cells | | CD8+ T cells | | FoxP3+ T cells | |
|---|---|---|---|---|---|---|
| | **Epithelium $\chi2$ (p-value)** | **Stroma $\chi2$ (p-value)** | **Epithelium $\chi2$ (p-value)** | **Stroma $\chi2$ (p-value)** | **Epithelium $\chi2$ (p-value)** | **Stroma $\chi2$ (p-value)** |
| **Age** | | | | | | |
| < 71 years | 0.216 | 0.009 | 0.076 | 0.076 | 2.479 | 1.451 |
| ≥ 71 years | (0.642) | (0.924) | (0.783) | (0.783) | (0.115) | (0.228) |
| **BMI** | | | | | | |
| Normal | 0.942 | 2.522 | 0.071 | 0.353 | 1.773 | 2.754 |
| Overweight/obese | (0.332) | (0.112) | (0.791) | (0.553) | (0.183) | (0.097) |
| **FIGO stage** | | | | | | |
| Stage I & II | 0.427 | 0.068 | 0.838 | 0.017 | 0.274 | 2.069 |
| Stage III & IV | (0.513) | (0.794) | (0.360) | (0.896) | (0.601) | (0.150) |
| **Tumor size** | | | | | | |
| ≤ 4cm | 0.115 | 0.004 | 0.547 | 3.770 | 0.004 | 0.004 |
| > 4cm | (0.735) | (0.951) | (0.460) | (0.052) | (0.951) | (0.951) |
| **Lymph node metastasis** | | | | | | |
| No | 0.211 | 0.004 | 1.255 | 0.109 | 0.109 | 1.559 |
| Yes | (0.646) | (0.949) | (0.263) | (0.741) | (0.741) | (0.212) |
| **HPV DNA** | | | | | | |
| Negative | 0.020 | 1.145 | 0.901 | 2.172 | 0.297 | 0.297 |
| Positive | (0.887) | (0.285) | (0.343) | (0.141) | (0.586) | (0.586) |
| **HPV mRNA ISH** | | | | | | |
| Negative | 1.479 | 0.228 | 0.628 | 0.812 | 0.628 | 0.628 |
| Positive | (0.224) | (0.633) | (0.428) | (0.367) | (0.428) | (0.428) |
| **p53/p16** | | | | | | |
| p53+/p16- | 1.398 | 1.987 | 2.814 | 2.226 | 1.381 | 1.638 |
| p53-/p16+ | (0.497) | (0.370) | (0.245) | (0.329) | (0.501) | (0.441) |
| p53-/p16- | | | | | | |
| **Recurrence of cancer** | | | | | | |
| No | 1.255 | 0.109 | 0.004 | 0.004 | 1.255 | 1.255 |
| Yes | (0.263) | (0.741) | (0.949) | (0.949) | (0.263) | (0.263) |
| **Death caused by VSCC** | | | | | | |
| No | 0.553 | 0.553 | 1.461 | 0.035 | **4.577** | 0.077 |
| Yes | (0.457) | (0.457) | (0.227) | (0.852) | **(0.032)** | (0.781) |

**Supplementary Table 3:** Relationship between epithelial and stromal infiltration of immune cells with clinicopathological features.

| Clinicopathological features | CD68+ macrophages | | CD14+ monocytes | |
|---|---|---|---|---|
| | **Epithelial $\chi2$ (p-value)** | **Stromal $\chi2$ (p-value)** | **Epithelial $\chi2$ (p-value)** | **Stromal $\chi2$ (p-value)** |
| **Age** | | | | |
| < 71 years | 3.079 | 3.079 | 0.416 | **8.206** |
| ≥ 71 years | (0.079) | (0.079) | (0.519) | **(0.004)** |
| **BMI** | | | | |
| normal | 0.406 | 0.575 | 1.888 | 1.011 |
| overweight/obese | (0.524) | (0.448) | (0.169) | (0.315) |
| **FIGO stage** | | | | |
| Stage I & II | **6.171** | 2.889 | 0.838 | 1.710 |
| Stage III & IV | **(0.013)** | (0.089) | (0.360) | (0.191) |
| **Tumor size** | | | | |
| ≤ 4cm | **5.486** | **5.486** | **5.486** | **7.523** |
| > 4cm | **(0.019)** | **(0.019)** | **(0.019)** | **(0.006)** |
| **Lymph node metastasis** | | | | |
| No | **7.289** | 2.296 | 0.109 | 2.296 |
| Yes | **(0.007)** | (0.130) | (0.741) | (0.130) |
| **HPV DNA** | | | | |
| Negative | **7.211** | **7.211** | 0.069 | **7.211** |
| Positive | **(0.007)** | **(0.007)** | (0.793) | **(0.007)** |
| **HPV mRNA ISH** | | | | |
| Negative | **4.716** | **6.734** | 0.228 | **4.716** |
| Positive | **(0.030)** | **(0.009)** | (0.633) | **(0.030)** |
| **p53/p16** | | | | |
| p53+/p16- | **6.873** | 4.851 | 1.268 | **6.274** |
| p53-/p16+ | **(0.032)** | (0.088) | (0.531) | **(0.043)** |
| p53-/p16- | | | | |
| **Recurrence of cancer** | | | | |
| No | 1.255 | 0.526 | 1.255 | 1.255 |
| Yes | (0.263) | (0.468) | (0.263) | (0.263) |
| **Death caused by VSCC** | | | | |
| No | 0.035 | 2.505 | 0.077 | 2.505 |
| Yes | (0.852) | (0.113) | (0.781) | (0.113) |

**Supplementary Table 4:** Univariate Cox regression analysis to determine the influence of clinicopathological features on progression free- and disease specific survival.

| Clinicopathological features | Progression free survival | | | Disease specific survival | | |
|---|---|---|---|---|---|---|
| | HR | (95% CI) | p-value | HR | (95% CI) | p-value |
| **Age**<br>< 71 years<br>≥ 71 years | **2.172** | **1.240-3.805** | **0.007** | 4.535 | 1.738-11.833 | **0.002** |
| **BMI**<br>Normal<br>Overweight and obese | **2.322** | **1.047-5.150** | **0.038** | 2.857 | 0.799-10.208 | 0.106 |
| **FIGO stage**<br>Stage I<br>Stage II<br>Stage III<br>Stage IV | 1.214 | 0.905-1.628 | 0.195 | 1.344 | 0.864-2.089 | 0.190 |
| **Tumor size**<br>≤ 4cm<br>> 4cm | 1.692 | 0.944-3.034 | 0.078 | **2.487** | **1.052-5.881** | **0.038** |
| **Lymph node metastasis**<br>No<br>Yes | 1.337 | 0.749-2.386 | 0.327 | 1.228 | 0.503-2.998 | 0.653 |
| **HPV DNA**<br>Negative<br>Positive | **0.188** | **0.080-0.442** | **<0.001** | **0.133** | **0.031-0.574** | **0.007** |
| **HPV mRNA ISH**<br>Negative<br>Positive | **0.144** | **0.052-0.402** | **<0.001** | **0.075** | **0.010-0.564** | **0.012** |
| **Recurrence of cancer**<br>No<br>Yes | - | - | - | **2.854** | **1.189-6.847** | **0.019** |
